# Diffusion MRI abnormalities localise the epileptogenic zone in paediatric drug-resistant epilepsy

**DOI:** 10.64898/2026.09.25.26363785

**Authors:** Damjan Veljanoski, Aswin Chari, Kiran K. Seunarine, Felice D’Arco, Kshitij Mankad, Zubair Tahir, Chris A. Clark, Martin M. Tisdall, Torsten Baldeweg, Rory J. Piper

## Abstract

**Objectives:** Seizure freedom follows surgery in 67% of children with drug-resistant epilepsy, but imaging biomarkers could improve patient selection, epileptogenic zone localisation, and outcome prediction. We assessed whether diffusion MRI abnormalities predicted seizure freedom when resected; aligned with stereo-EEG seizure-onset and interictal zones; distinguished patients with/without an identified onset zone; and localised the epileptogenic zone in MRI-negative cases.

**Methods:** We retrospectively studied children undergoing focal resection (n=110) or stereo-EEG (n=61) between 2015–2023, alongside healthy controls aged 6–18 (n=69), using identical 3T MRI diffusion protocols. Age- and sex-adjusted normative models generated mean diffusivity (MD) and fractional anisotropy (FA) voxelwise z-score maps to identify dominant abnormalities. In resection patients, abnormality–resection overlap was related to ≥12-month Engel outcome. In stereo-EEG patients, z-scores were compared across seizure-onset, interictal, and uninvolved grey-matter contacts. In lesion-negative patients, abnormalities were compared with PET hypometabolism and stereo-EEG findings.

**Results:** Greater resection of MD abnormalities predicted seizure freedom, especially with overlap >0 (mean differences 17%, 23%; *P*_perm_=0.0086, <0.001), and higher odds of Engel I (OR 1.31/10%; AUC 0.723; Youden threshold 36%). FA showed similar associations. Seizure-onset contacts had higher MD and lower FA z-scores than uninvolved grey matter (+0.65, −0.18; *P*_perm_<0.001, =0.001), with interictal contacts intermediate. Abnormalities were more widespread and contralateral in patients without an identified onset zone. In lesion-negative patients, abnormalities co-localised with PET (20/28 fully, 6/28 partially) and stereo-EEG (17/24 fully, 3/24 partially).

**Interpretation:** Diffusion MRI abnormalities aligned with invasive electrophysiology, predicted seizure freedom when resected, and co-localized with PET/stereo-EEG, supporting an important role in presurgical evaluation.

## Introduction

67% of children undergoing resective surgery for drug-resistant epilepsy (DRE) are seizure free,^1^ but seizure freedom rates are lower in patients with non-lesional epilepsy or incomplete resection of the epileptogenic zone (EZ).^2^ Stereo-EEG is now integral to multimodal pre-operative epilepsy evaluation and has improved localization, but is invasive and costly.^3^ PET imaging is another adopted method for detecting the EZ, but exposes the child to additional radiation. Therefore, there remains a persistent, unmet need for reliable, non-invasive pre-operative imaging biomarkers that can improve surgical candidate selection and accurately delineate the EZ.

Diffusion MRI (dMRI) provides voxelwise measures of white matter (WM) integrity^4^, and the two most commonly studied metrics are mean diffusivity (MD) and fractional anisotropy (FA).^5,6^ Increased MD represents a loss of tissue microstructure whilst decreased FA is related to loss of WM fiber coherence, reduced axonal density, or both.^7–9^ Prior studies have shown that increased MD often co-localized with irritative zone stereo-EEG contacts^10^ and that combining dMRI with FDG-PET improves localization in MRI-negative epilepsy, particularly in extra-temporal cases.^11^ More recently, partial resection of dMRI abnormalities has been associated with seizure freedom in adults.^12^ However, the earlier studies used 1.5 Tesla MRI systems and both earlier and more recent studies have been conducted almost exclusively in adult cohorts, limiting the direct applicability of these findings to children, in whom diffusion metrics change with brain maturation.^13,14^

The objective of this study was to evaluate the diagnostic and prognostic potential of normative dMRI in children with focal DRE. Firstly, we characterized patient-specific dMRI-derived abnormalities and tested the primary hypothesis in a resective surgery cohort that a greater proportion of the dominant dMRI abnormality resected is associated with postoperative seizure freedom. Secondly, we examined diffusion-derived metrics at stereo-EEG contact locations, comparing seizure onset zone (SOZ) contacts, interictal zone (IZ) contacts and remaining gray matter (GM) contacts. Thirdly, we compared group-level diffusion abnormalities between patients in whom stereo-EEG identified an SOZ and those in whom no SOZ was identified, as these groups may represent focal and more distributed patterns of disease, respectively. Finally, we assessed the utility of this method in clinically challenging patients with MRI lesion-negative epilepsy by determining whether diffusion abnormalities are spatially concordant with PET hypometabolism and stereo-EEG findings.

## Materials and methods

### Standard Protocol Approvals, Registrations, and Patient Consent

This retrospective observational study was approved by the Research Governance Office of Great Ormond Street Hospital (ID: 23NP01).

### Patient and control cohorts

This study comprised two surgical cohorts: those undergoing resection and stereo-EEG. Data was included for patients aged 7-18 years (+/-11 months) to match the age range of the healthy controls. All patients underwent surgery at GOSH following discussion in an epilepsy surgery multidisciplinary team meeting. Consistent with prior methodology,^15^ across all cohorts, patients were excluded if MRI was acquired using a different scanner or protocol (to ensure imaging compatibility and data homogeneity), or if they had tuberous sclerosis complex, prior resective surgery, or inadequate preprocessing (Supplementary Table 3). The resective cohort included patients with DRE undergoing resection between 2015–2023 who had a post-operative volumetric T1-weighted MRI sequence (to delineate the resection volume). The stereo-EEG cohort included patients with presumed focal DRE undergoing stereo-EEG between 2015–2023. Patients were classified as seizure-free (SF; Engel I) or not seizure-free (NSF; Engel II–IV) at last follow-up, ≥12 months post-operatively. Controls were healthy children from our previous neuroimaging studies^16–18^ scanned using the same dMRI protocol and scanner as the patient cohorts.

### MRI acquisition and processing

MRI data were acquired on a single 3T scanner using a multi-shell diffusion protocol with consistent acquisition parameters across all control and patient cohorts. Resection masks were manually delineated and transformed between structural, diffusion, and template spaces (fig. 1A). Structural and diffusion data were processed using a standardized pipeline (*micapipe*, ^19^ and all images were registered to a population-derived template (Figure 1B). Detailed acquisition parameters and MRI preprocessing steps are provided in the Supplementary Methods Section 1.

**Figure 1.**
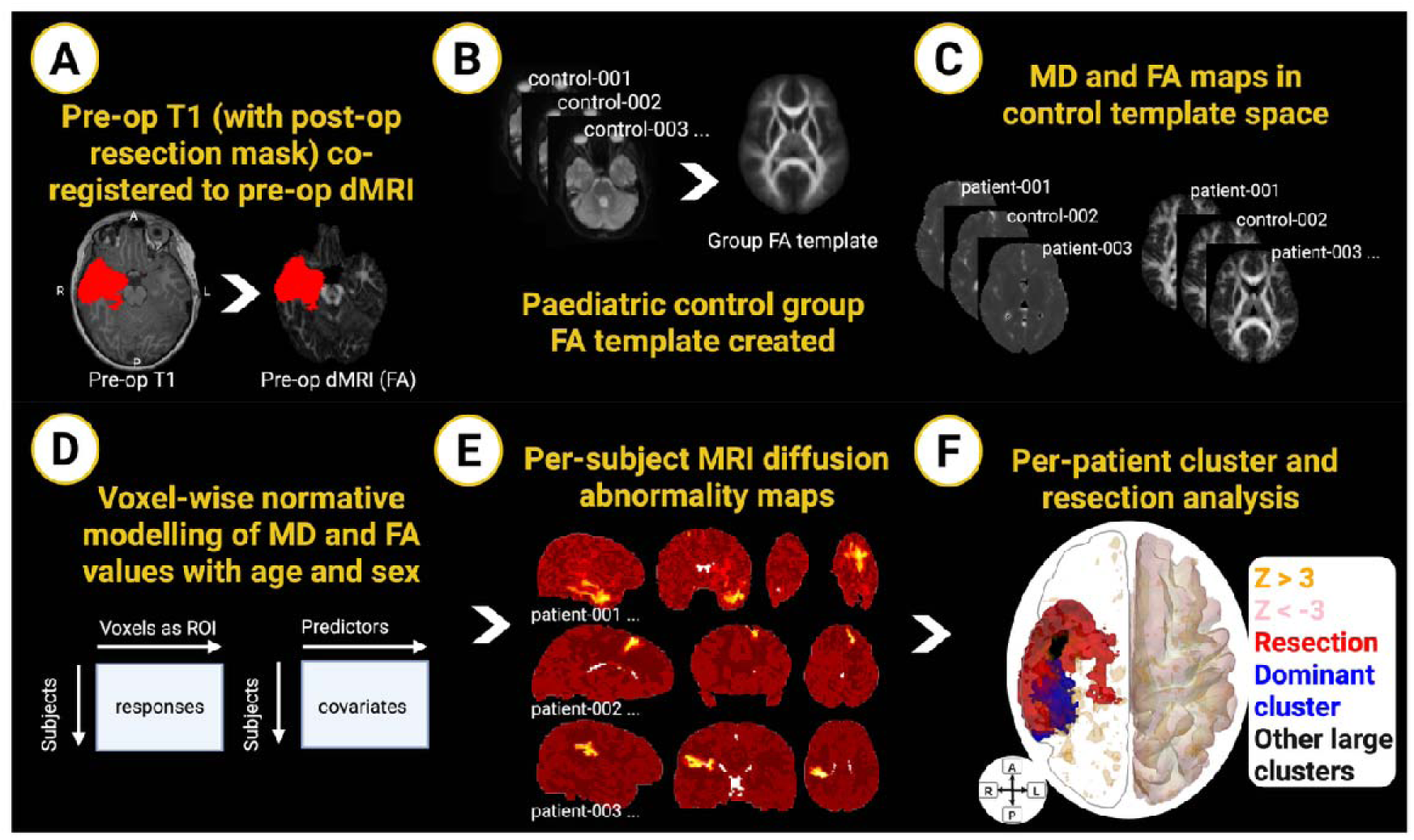
Summary of registration steps, normative modelling and clustering. **(A)** Co-registration of preoperative T1, diffusion and (where applicable) postoperative T1 scans with resection masks (red region). **(B)** Creation of a control group T1 and FA template. **(C)** Registration of FA and MD maps to the FA template. **(D)** Voxel-wise normative modelling of MD and FA maps adjusting for age and sex using the PCNToolkit. **(E)** Per-patient MD and FA diffusion abnormality maps (MD shown in this figure). **(F)** Per-patient whole-brain cluster and resection visualization.

### Normative modelling

Voxelwise normative modelling was used to identify patient-specific MD and FA abnormalities relative to healthy controls. We first confirmed expected age-related decreases in MD and increases in FA in a representative right temporal region of interest in all patients and controls (Supplementary Fig. 1).^13,14^

Three approaches to standardizing MD and FA values were compared: conventional z-scoring, age- and sex-adjusted generalized linear modelling (GLM), and Bayesian linear regression (BLR) using PCNtoolkit^20^. As GLM and BLR produced highly concordant voxelwise maps in an initial comparison (Supplementary Fig. 2), the latter approach (Fig. 1D) was used for all downstream analyses because it also supports scalable adjustment for multi-site and scanner-related effects.

Control and patient MD and FA maps were restricted to a group intersection brain mask and stacked into 4D NIfTI files. Age at preoperative scan and sex were included as covariates. The BLR model was fitted voxelwise in controls only, after covariate standardization, and then applied to patient data to generate individual standardized deviation (z-score) maps for MD and FA (Fig. 1E).

### Abnormal diffusion cluster selection

We first systematically optimized clustering parameters to maximize anatomical concordance between diffusion-derived abnormalities and surgical resection sites in a subset of patients that were seizure free after lesionectomy. Clusters in individual z-scored MD and FA maps (Fig. 1E) were enhanced using an empirical grid search over percentile thresholds (95, 99), voxel connectivity (6, 18, 26), Gaussian smoothing (σ=0–4 voxels), and threshold-free cluster enhancement (TFCE) parameters (H=1–4; E=0.1–4)^21^. For each parameter combination, the full cluster enhancement pipeline was applied separately to MD and FA z-maps, and performance was quantified as the mean percentage overlap between resection masks and diffusion abnormalities in SF patients following lesionectomy because it can be assumed for this cohort that the putative SOZ was correctly identified and successfully targeted. The final parameter set (PCTL=99, 26-connectivity, σ=1 voxel, H=2, E=0.5) was selected based on consistently high overlap among the top-performing combinations. Consistent with prior work^10–12,22–24^, subsequent analyses focused on positive MD and negative FA z-scores.

Using these optimized parameters, z-maps were smoothed (σ=1 voxel) and TFCE parameters were applied (Fig. 2A–B). Negative z-scores were sign-inverted prior to TFCE and restored afterwards using identical settings. Voxels exceeding the 99th percentile of TFCE values were clustered using 26-voxel connectivity, and clusters were ranked by TFCE mass (defined as the sum of TFCE values within the cluster). A data-driven changepoint approach based on a two-segment piecewise linear model identified a breakpoint in the ranked TFCE-mass distribution, with clusters above this threshold retained as ‘large’ clusters (Fig. 1F). We also explored thresholding the MD z-maps at z>3 only without further optimization and defining clusters using 26-voxel connectivity with a minimum extent of 1 voxel, before identifying large clusters using the change-point analysis (Supplementary Fig. 15).

**Figure 2.**
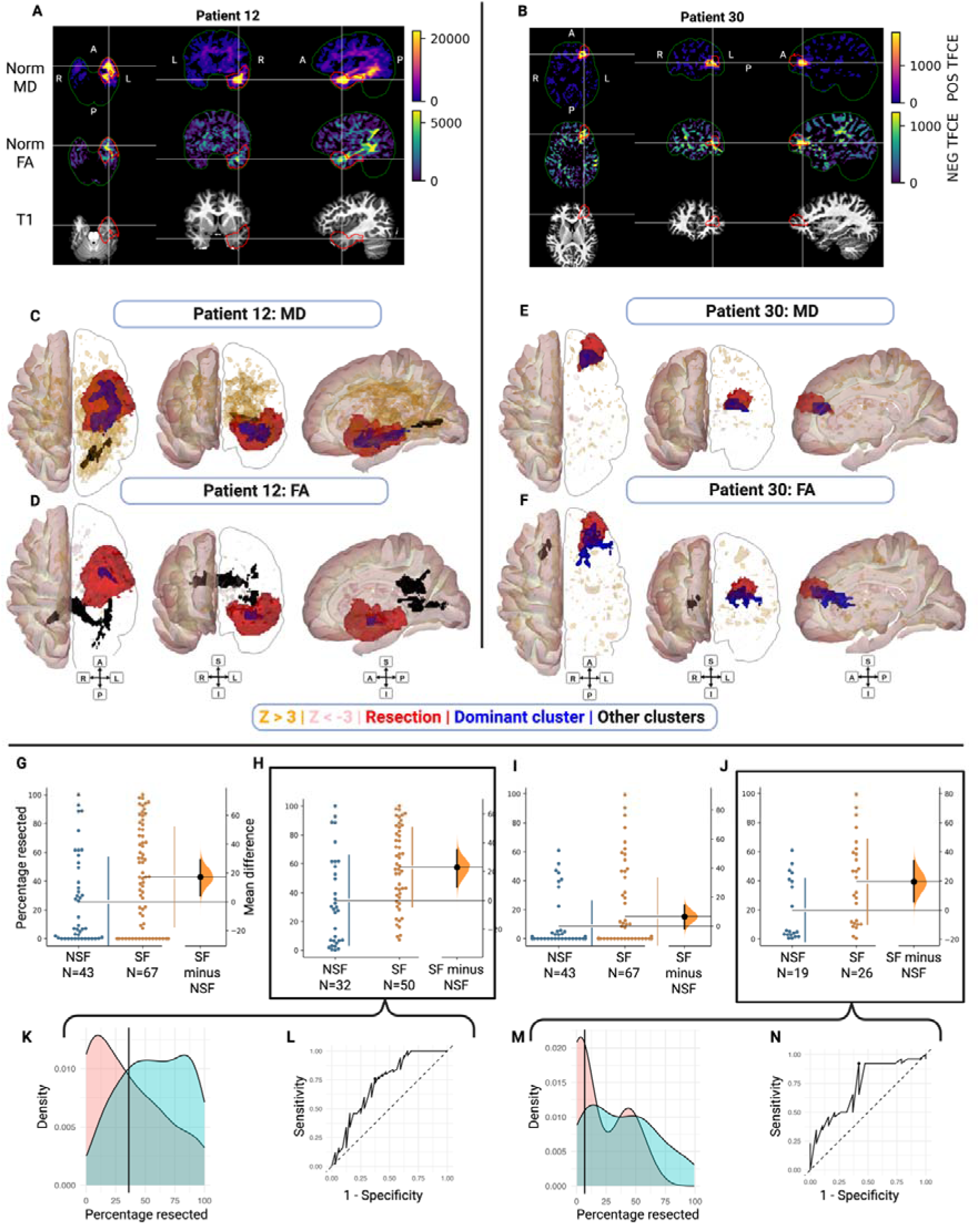
Top panel: Two representative patient cases. Bottom panels: Extent of resection of abnormal diffusion MD and FA clusters and association with seizure freedom. **(A)** unthresholded TFCE normative MD map (top panel: using positive z-values, reflecting elevated diffusivity relative to controls), negative FA (middle panel: using negative z-values, reflecting reduced anisotropy relative to controls), T1 (bottom panel) maps in radiological orientation for patient 12, MRI showed ill-defined loss of gray-white matter differentiation in the left temporal lobe, who underwent left temporal lobectomy (red outline) for a DNET tumor, subsequently SF. **(B)** Analogous maps for patient 30, who underwent left frontal lesionectomy for an FCD IIId, subsequently SF. **(C)** Three-dimensional reconstructions of the MD-derived map for patient 12, showing abnormal clusters (blue; dominant cluster, black; other large clusters, voxels with z-score>3; orange, voxels with z-score<-3; pink). **(D)** FA-derived map, analogous to **(C)** for patient 12. **(E, F)** MD- and FA-maps for patient 30, analogous to **(C, D)**. **(G, H)** Gardner-Altman plots of the mean difference in the percentage resected of the dominant MD cluster between SF and NSF patients. SF and NSF groups are plotted on the left axes; the mean difference is plotted on a floating axis on the right as a bootstrap sampling distribution. The mean difference is depicted as a dot; the 95% confidence interval is indicated by the ends of the vertical error bar. **(G)** Depicts all patients, including those with none of the dominant cluster resected whereas **(H)** depicts only patients with >0% of the dominant cluster resected. **(K)** Density plot of the percentage resected of the dominant MD cluster (for patients with >0% resected) versus the probability density with the vertical line denoting the Youden optimal threshold from the ROC analysis. **(L)** ROC curve shows sensitivity versus 1 − specificity for predicting seizure freedom based on the percentage resected of the dominant MD-derived cluster for patients with >0% resection. The diagonal line indicates chance performance, and the marked point corresponds to the Youden-optimal cut-point. AUC summarizes discriminative ability. **(I, J, M, N)** are for the FA-derived dominant cluster, analogous to **(G, H, K, L)**.

To quantify the relationship between surgical resections and abnormal diffusion clusters, we calculated the percentage overlap between each patient’s resection mask and their dominant diffusion abnormality (Fig. 2C–F). The dominant abnormality was defined as the largest retained cluster (by TFCE mass) localizing to the resection lobe, or, if no such cluster was present, the ipsilateral hemisphere. By guiding cluster selection according to the resection lobe, which served as a surrogate for the presurgical hypothesized SOZ lobe, we simulated how a clinician would prioritize abnormalities during preoperative localization. We characterized the anatomical distribution (deep GM, WM or cortical GM) of each patient’s dominant diffusion abnormality in template space, stratified by outcome.

### Group MD and FA maps

To characterize group-level spatial patterns of diffusion abnormalities we generated aggregate voxelwise maps across stereo-EEG patients were stratified according to whether the SOZ was identified or not (Supplementary Methods: Section 2).

### Stereo-EEG electrode localization and registration

Stereo-EEG electrode contacts were localized and mapped into the same diffusion template space for quantitative comparison. Stereo-EEG electrode contacts were localized using a CT scan, identified using *SEEG assistant*,^25^ assigned voxel spaces and registered to the preoperative T1 scan using *reg_aladin*.^26,27^ The preoperative T1 scans were registered to the control group FA template using *reg_aladin*^26^ and the affine transforms were used to resample and dilate the electrode contacts as crosshairs into the diffusion space (Fig. 3A, 3D). Electrode contacts were designated as GM, seizure onset or main interictal discharge contacts by the consultant neurophysiologist at the post-implantation stereo-EEG multidisciplinary meeting. Absolute z-scores, including positive and negative values, were extracted from whole-brain MD and FA maps for each contact crosshair before calculating per-patient mean GM, seizure onset and interictal z-scores. To avoid overlapping electrode contact designations, contacts were assigned using a hierarchical exclusivity rule: seizure onset>interictal>GM. Per-patient mean z-scores were then calculated separately for the exclusive contact groups.

**Figure 3.**
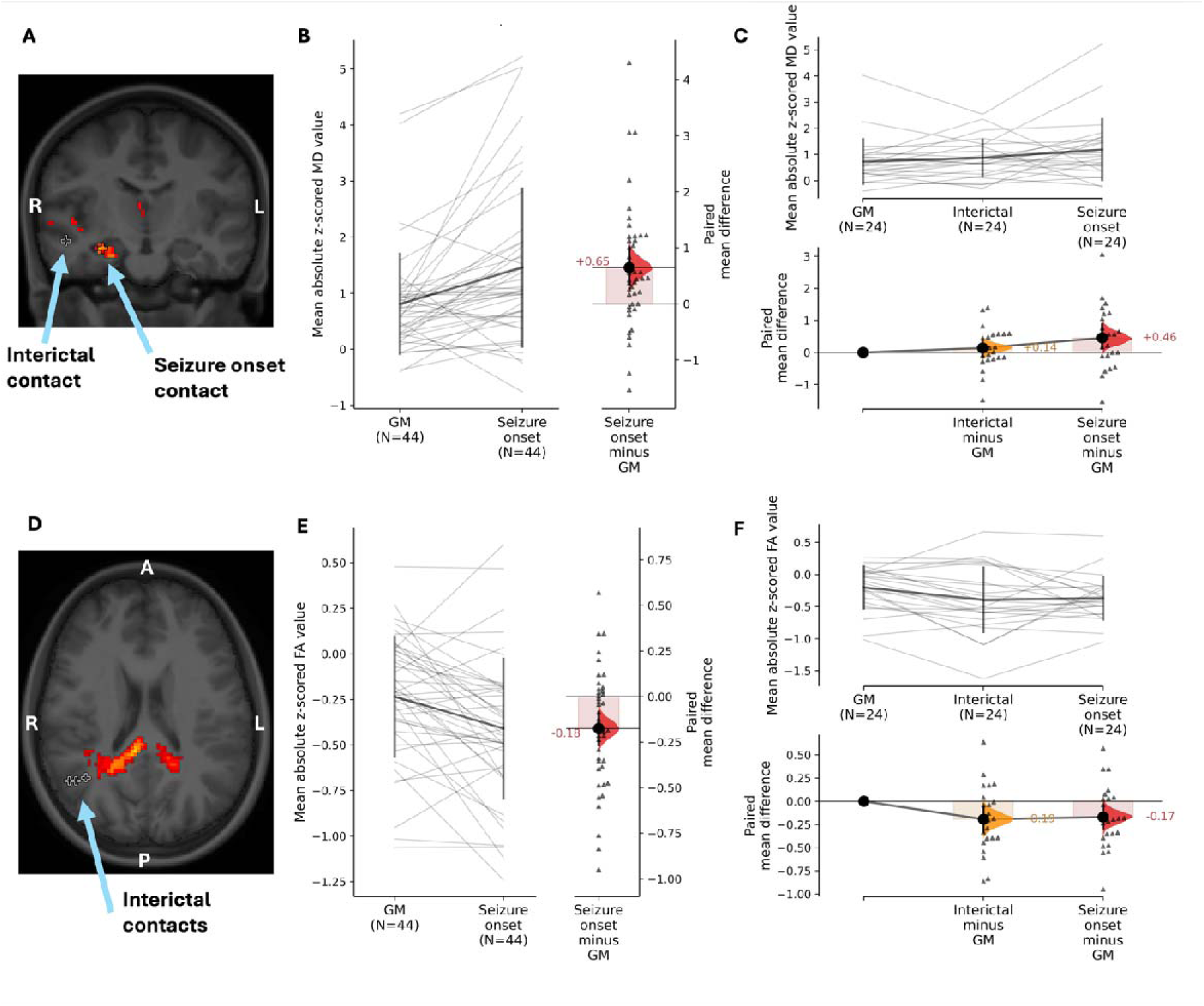
Paired estimation plots of MD and FA values across GM, interictal and seizure onset contacts. **(A)** MD abnormality shown relative to interictal and seizure onset contacts. **(B)** Paired estimation plot comparing mean z-scored MD values (positive and negative) between GM contacts (excluding the seizure onset contacts) and the seizure onset contacts (N=44). Thin gray lines represent paired patient-level values. The right panel shows the paired mean difference (seizure onset − GM) with bootstrap distribution, point estimate (black dot), and 95% bias-corrected and accelerated (BCa) confidence interval. **(C)** Repeated-measures estimation plot of MD across GM, interictal and seizure onset contacts for patients with complete triplet data (N=24). The lower panel shows paired mean differences relative to GM (interictal – GM; seizure onset − GM) with bootstrap distributions and 95% BCa confidence intervals. **(D)** FA abnormality shown relative to interictal and seizure onset contacts. **(E)** Paired estimation plot comparing mean absolute z-scored FA values between GM and seizure onset (N=44), displayed in the same format as in panel A. **(F)** Repeated-measures estimation plot of FA across GM, interictal, and seizure onset contacts (N=24), with paired mean differences relative to GM shown in the lower panel, analogous to panel **(C)**.

### Concordance of diffusion MRI abnormalities with PET hypometabolism and stereo-EEG findings

We examined the unthresholded TFCE MD (positive) and FA (negative) maps of patients with extratemporal and temporal MRI lesion-negative epilepsy (Supplementary Figs. 9-12). For PET concordance, the spatial distribution of diffusion abnormalities was compared with the region of PET hypometabolism as defined by the reporting neuroradiologist. For stereo-EEG concordance, diffusion abnormalities were compared with the spatial distribution of SOZ contacts in patients with an identified SOZ; in patients without an identified SOZ, diffusion abnormalities were compared with the regions implicated in the stereo-EEG clinical report. Findings were classified as concordant when diffusion abnormalities co-localized regionally with the PET or stereo-EEG findings, and as partially concordant when there was only partial spatial overlap (Supplementary Tables 1-2).

### Statistical analyses

Statistical analyses and figure preparations were performed using Rstudio version 2025.05.1+513, bioRender (using R version 4.2.2), Python and the estimation statistics web app.^28^ Control and patient cohorts were compared for differences in demographic, SOZ and outcome using non-parametric Mann Whitney tests, with significance defined as *P*<0.05. See Supplementary Materials Section 3, Statistical methods, for details of the statistical analyses—including linear regression models, bootstrap-based estimation statistics, ROC curve and AUC analyses, logistic regression modelling, and stereo-EEG-based paired comparisons.

## Results

### Patient and control cohort characteristics

Cohort details are summarized in Table 1. The resective cohorts (n=110; 61% SF, 39% NSF) showed comparable age at scan, age at surgery, epilepsy duration, and follow-up across outcome groups; most underwent temporal or frontal resections, with tumor, focal cortical dysplasia (FCD), and hippocampal sclerosis (HS) as the predominant pathologies. The stereo-EEG cohort (n=61) had similar ages at MRI and implantation regardless of SOZ identification (identified in 67%); 20 later underwent resection and two had repeat implantations.

**Table 1.**
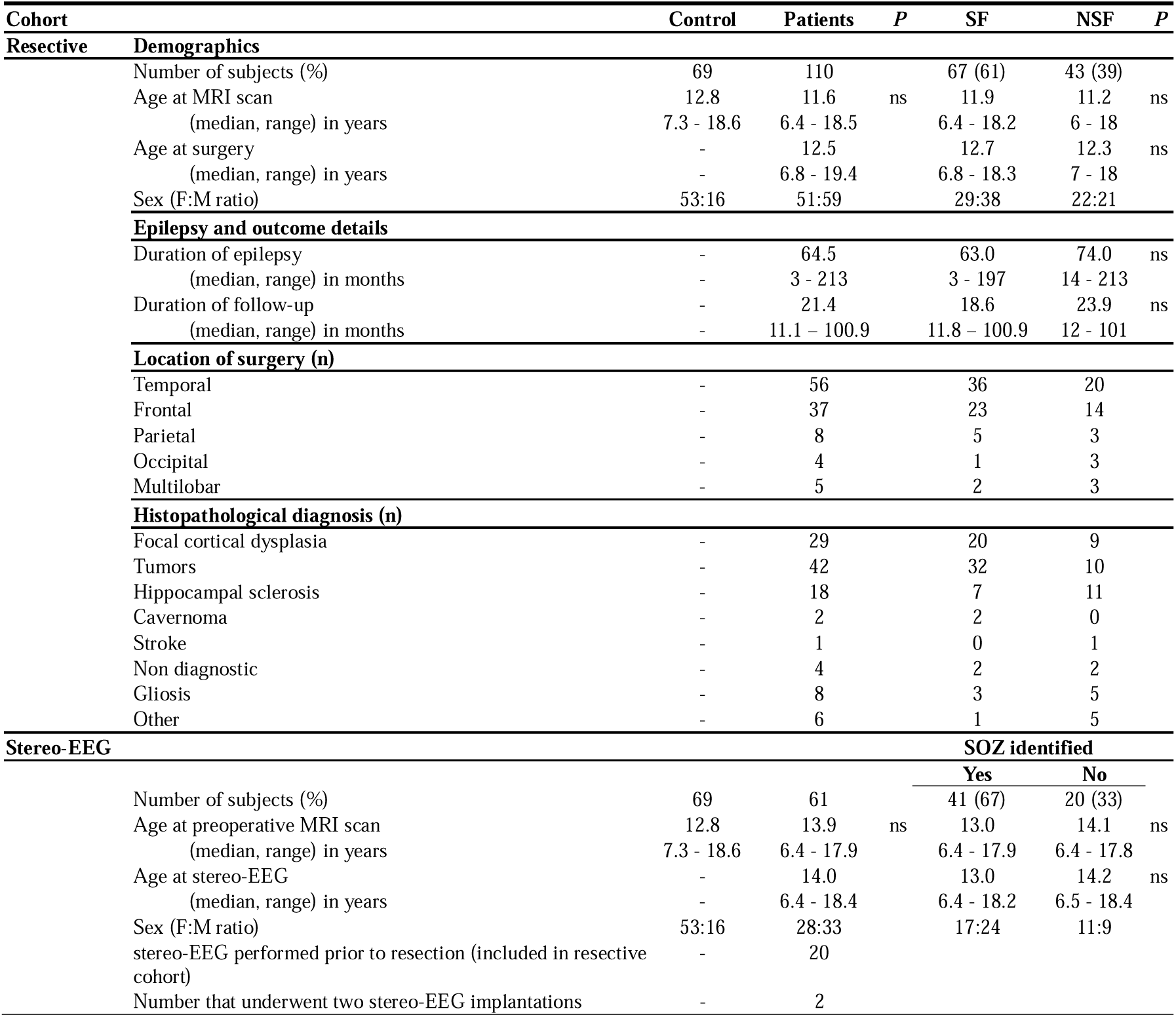
Patient and control cohort characteristics. *P*-value: ns if *P*≥0.05. Stereo-EEG; stereoelectroencephalography, SF; seizure-free, NSF; not seizure-free, SOZ; seizure onset zone.

| Cohort |  | Control | Patients | P | SF | NSF | P |
| --- | --- | --- | --- | --- | --- | --- | --- |
| **Resective** | **Demographics** |  |  |  |  |  |  |
|  | Number of subjects (%) | 69 | 110 |  | 67 (61) | 43 (39) |  |
|  | Age at MRI scan | 12.8 | 11.6 | ns | 11.9 | 11.2 | ns |
|  | (median, range) in years | 7.3 - 18.6 | 6.4 - 18.5 |  | 6.4 - 18.2 | 6 - 18 |  |
|  | Age at surgery | - | 12.5 |  | 12.7 | 12.3 | ns |
|  | (median, range) in years | - | 6.8 - 19.4 |  | 6.8 - 18.3 | 7 - 18 |  |
|  | Sex (F:M ratio) | 53:16 | 51:59 |  | 29:38 | 22:21 |  |
|  | **Epilepsy and outcome details** |  |  |  |  |  |  |
|  | Duration of epilepsy | - | 64.5 |  | 63.0 | 74.0 | ns |
|  | (median, range) in months | - | 3 - 213 |  | 3 - 197 | 14 - 213 |  |
|  | Duration of follow-up | - | 21.4 |  | 18.6 | 23.9 | ns |
|  | (median, range) in months | - | 11.1 - 100.9 |  | 11.8 - 100.9 | 12 - 101 |  |
|  | **Location of surgery (n)** |  |  |  |  |  |  |
|  | Temporal | - | 56 |  | 36 | 20 |  |
|  | Frontal | - | 37 |  | 23 | 14 |  |
|  | Parietal | - | 8 |  | 5 | 3 |  |
|  | Occipital | - | 4 |  | 1 | 3 |  |
|  | Multilobar | - | 5 |  | 2 | 3 |  |
|  | **Histopathological diagnosis (n)** |  |  |  |  |  |  |
|  | Focal cortical dysplasia | - | 29 |  | 20 | 9 |  |
|  | Tumors | - | 42 |  | 32 | 10 |  |
|  | Hippocampal sclerosis | - | 18 |  | 7 | 11 |  |
|  | Cavernoma | - | 2 |  | 2 | 0 |  |
|  | Stroke | - | 1 |  | 0 | 1 |  |
|  | Non diagnostic | - | 4 |  | 2 | 2 |  |
|  | Gliosis | - | 8 |  | 3 | 5 |  |
|  | Other | - | 6 |  | 1 | 5 |  |
| **Stereo-EEG** |  |  |  |  | **SOZ identified** |  |  |
|  |  |  |  |  | **Yes** | **No** |  |
|  | Number of subjects (%) | 69 | 61 |  | 41 (67) | 20 (33) |  |
|  | Age at preoperative MRI scan | 12.8 | 13.9 | ns | 13.0 | 14.1 | ns |
|  | (median, range) in years | 7.3 - 18.6 | 6.4 - 17.9 |  | 6.4 - 17.9 | 6.4 - 17.8 |  |
|  | Age at stereo-EEG | - | 14.0 |  | 13.0 | 14.2 | ns |
|  | (median, range) in years | - | 6.4 - 18.4 |  | 6.4 - 18.2 | 6.5 - 18.4 |  |
|  | Sex (F:M ratio) | 53:16 | 28:33 |  | 17:24 | 11:9 |  |
|  | stereo-EEG performed prior to resection (included in resective cohort) | - | 20 |  |  |  |  |
|  | Number that underwent two stereo-EEG implantations | - | 2 |  |  |  |  |

### A greater extent of resection of diffusion MRI abnormalities is associated with post-surgical seizure freedom

82 of 110 patients had overlap between the resection mask and the dominant positive MD abnormality, and 45 had overlap between the resection mask and the dominant negative FA abnormality selected based on the involved lobe (Fig. 2A–F). Dominant MD and FA abnormalities were mostly in the WM, followed by the cortical GM and the deep GM, with similar proportions across SF and NSF patients (Supplementary Fig. 3A-B).

For positive MD–derived clusters, the proportion of the dominant abnormality resected was greater in SF (mean 43%) than NSF (mean 26%) patients. Across all patients, the mean difference between SF and NSF patients was 17.3% (CI 4.25, 29.2; *P*_perm_=0.0086; Fig. 2G). The percentage resected was greater in patients who underwent lobectomy compared with lesionectomy (two-way ANOVA, F(1,106)=7.614) however there was no interaction between operation type and outcome (F(1,106)=0.147), indicating that the association between percentage resected and seizure freedom did not differ by the operation type (Supplementary Fig. 4). When restricting the analysis to patients with >0% of the MD cluster resected, the mean difference was 23% (CI 9.15, 35.1; *P*_perm_=0.0008; Fig. 2H). For negative FA–derived clusters, the overall difference between SF and NSF patients was smaller (6.54%, CI −1.97, 14.5; *P*_perm_=0.149; Fig. 2I). However, in the subgroup with >0% FA cluster resected, SF patients had a greater proportion resected (19.6%, CI 5.75, 33.8; *P*_perm_=0.019; Fig. 2J).

Greater resection of the dominant positive MD abnormality (in patients with >0% resected) was associated with higher odds of seizure freedom (OR 1.31 per 10% increase, CI 1.10, 1.56; *P*=0.003). ROC analysis (Figs. 2K-L) indicated a moderate ability of the percentage of abnormal tissue resected to distinguish between SF and NSF patients (AUC 0.723, CI 0.604, 0.843). The Youden-optimal threshold was 36% resection (sensitivity 0.76, specificity 0.63), suggesting that resection of approximately one-third of the dominant abnormality provided the best statistical separation between outcome groups, although the precise value was widely variable on bootstrap resampling (median 33.3%, percentile interval 7.2–63.5). Similarly, greater resection of the dominant negative FA abnormality (in patients with >0% resected) was associated with higher odds of seizure freedom (OR 1.35, CI 1.06, 1.72; *P*=0.015) and comparable moderate discrimination (AUC 0.731, CI 0.578, 0.884; Figs. 2M-N). The Youden-optimal threshold was 6.8% (sensitivity 0.92, specificity 0.58), indicating that even relatively small proportions of the FA-defined abnormality contributed to outcome separation, although bootstrap analyses again demonstrated variability in the exact point (median 6.84%, percentile interval 6.29 to 53.09).

A substantial proportion of lesionectomy patients did not have any of their dominant diffusion abnormality resected (Supplementary Fig. 4). Therefore, we also examined the relationship between histology (FCD, HS and tumor) and percentage resected, stratified by outcome and found that this observed effect was largely contributed to by patients with FCDs (Supplementary Fig. 5). Of the 29 patients with FCDs, there were 8 who did not have any amount of their dominant diffusion abnormality resected across the MD or FA maps (discussed later).

### Diffusion MRI metrics are concordant with the seizure onset zone and interictal zone in patients undergoing stereo-EEG

Across 44 patients, MD was higher at seizure onset contacts than at remaining GM stereo-EEG contacts (Fig. 3B; *P*_perm_=0.0004). FA was lower at seizure onset contacts than at GM contacts (Fig. 3E; *P*_perm_=0.001). In the subset of patients with distinct interictal and seizure onset contacts (n=24), MD was higher at seizure onset contacts than at GM contacts (Fig. 3C; *P*_perm_=0.031). MD did not differ between interictal and GM stereo-EEG contacts (Fig. 3C; *P*_perm_=0.300). FA was lower at interictal contacts than at GM contacts (Fig. 3F; *P*_perm_=0.0176). FA was also lower at seizure onset contacts than at GM contacts (Fig. 3F; *P*_perm_=0.0212).

### Aggregate voxel-wise group maps reveal differences between SOZ-found versus SOZ-not-found stereo-EEG patients

For the stereo-EEG cohort, the between-group mean MD difference map indicated higher mean z-scores bilaterally in patients without an identified SOZ, most prominently within the temporal and frontal lobes contralateral to the presumed SOZ (Supplementary Fig. 7G). The corresponding median difference map (Supplementary Fig. 7H) showed a similar spatial distribution, suggesting that the observed group differences are not primarily driven by outlier patients. The fraction suprathreshold map (Supplementary Fig. 7I) demonstrated a higher prevalence of z≥2 abnormalities in patients without an identified SOZ, again predominantly contralateral. The corresponding stereo-EEG group FA maps were broadly concordant, though the differences were less pronounced (Supplementary Fig. 8).

### Diffusion abnormalities are spatially concordant with PET hypometabolism and stereo-EEG findings

Diffusion abnormalities were spatially concordant with PET hypometabolism (Fig. 4C) and, in patients who underwent stereo-EEG, were spatially concordant with the stereo-EEG findings (Fig. 4D). Even without thresholding, the maps frequently demonstrated clear hemispheric asymmetry with evident lateralization and lobar localization (Figs. 4A-B). Out of 28 lesion-negative patients who underwent PET, diffusion abnormalities were concordant with PET in 20 patients, partially concordant in 6 patients and non-concordant in 2 patients (Fig. 4C; Supplementary Figs. 9-12; Supplementary Tables 1-2). Out of 24 patients who underwent stereo-EEG, diffusion abnormalities were spatially concordant with the stereo-EEG findings in 17 patients, partially concordant in 3 patients and non-concordant in 4 patients (Fig. 4D; Supplementary Figs. 9-12; Supplementary Tables 1-2).

**Figure 4.**
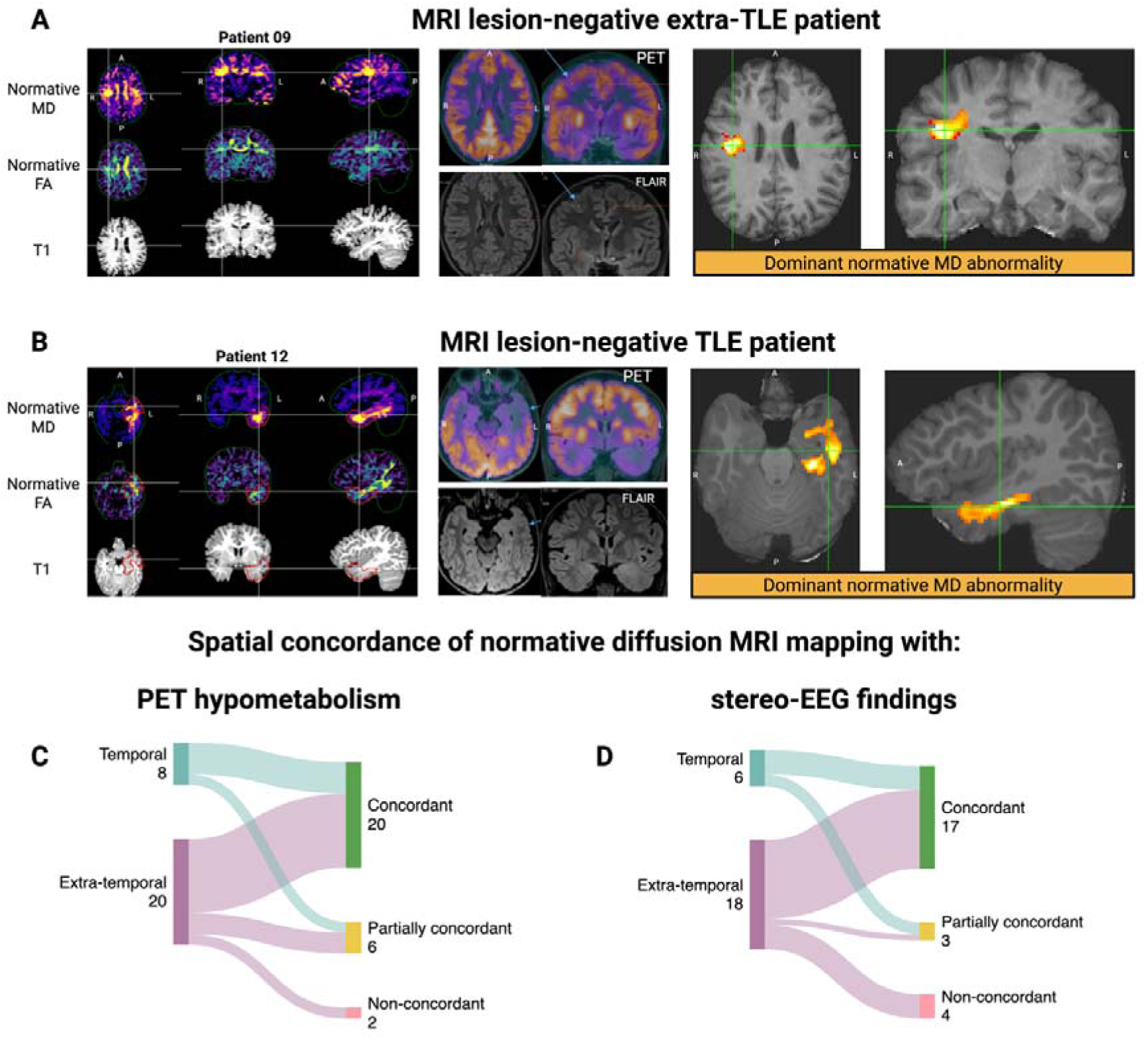
Per-patient normative diffusion abnormalities in radiological orientation for two MRI lesion-negative patients with right-sided extra-TLE (A) and left-sided TLE (B), with PET and FLAIR images for comparison. Left top panel: unthresholded TFCE normative MD map (using positive z-values, reflecting elevated diffusivity relative to controls). Left middle panel: unthresholded TFCE normative FA map (using negative z-values, reflecting reduced anisotropy relative to controls). Left bottom panel: Linearly aligned T1 scan in diffusion template space. Black crosshairs: SOZ-designated stereo-EEG contacts (where applicable). Red outline: resection masks (where applicable). Middle panel: PET scan with PET and FLAIR (labelled). Right panel: dominant normative MD abnormality. **(A)** Patient 09; with no definite lesion on MRI but right superior frontal gyrus showed transversely oriented sulcation anteriorly with mild gyral expansion and blurring of the gray-white junction. PET was suggestive of right frontal onset, probably within the middle frontal gyrus (MFG). Stereo-EEG found a widespread irritative zone with electrographic seizures in MFG, superior frontal gyrus, and orbitofrontal region; SOZ not found. The patient subsequently underwent vagus nerve stimulation achieving seizure freedom. **(B)** Patient 12; with ill-defined loss of gray-white matter differentiation in the left temporal lobe. PET showed left temporal hypometabolism. They underwent left temporal lobectomy (histology: DNET (WHO 1), achieving seizure freedom. **(C, D)** Sankey diagrams showing the spatial concordance of normative diffusion MRI mapping abnormalities with PET hypometabolism and stereo-EEG findings (Supplementary Figs. 24-27 and Supplementary Tables 1 and 2). **(C)** Concordance across MRI lesion-negative patients who had PET images available for review and those with PET reports only. **(D)** concordance across MRI lesion-negative patients who underwent stereo-EEG.

## Discussion

This study supports the role of dMRI in localizing the EZ in children with drug-resistant epilepsy. Critically, this approach demonstrates abnormalities concordant with the EZ not found on routine MRI (lesion negative cases) and is a promising non-invasive adjunct that could be utilized in presurgical evaluation. Across resective surgery and stereo-EEG cohorts, we demonstrate several key findings. SF patients had a greater proportion of the dominant diffusion abnormality resected than NSF patients, with increasing resection associated with higher odds of seizure freedom. Diffusion abnormalities were concordant with invasive electrophysiology findings, while group-level analyses showed more widespread, bilateral abnormalities in patients without an identified SOZ. In MRI lesion-negative cases, diffusion abnormalities were concordant with PET hypometabolism and stereo-EEG findings. Collectively, these findings suggest normative dMRI can improve clinical decision-making by directing attention to subtle abnormalities, particularly in MRI-negative patients, and may reduce the need for additional investigations such as PET. It may support epilepsy surgery pathways (Fig. 5) by guiding stereo-EEG implantation, informing candidate selection, and predicting postoperative outcome, while also identifying children with multifocal or diffuse disease unlikely to benefit from further invasive work-up.

**Figure 5.**
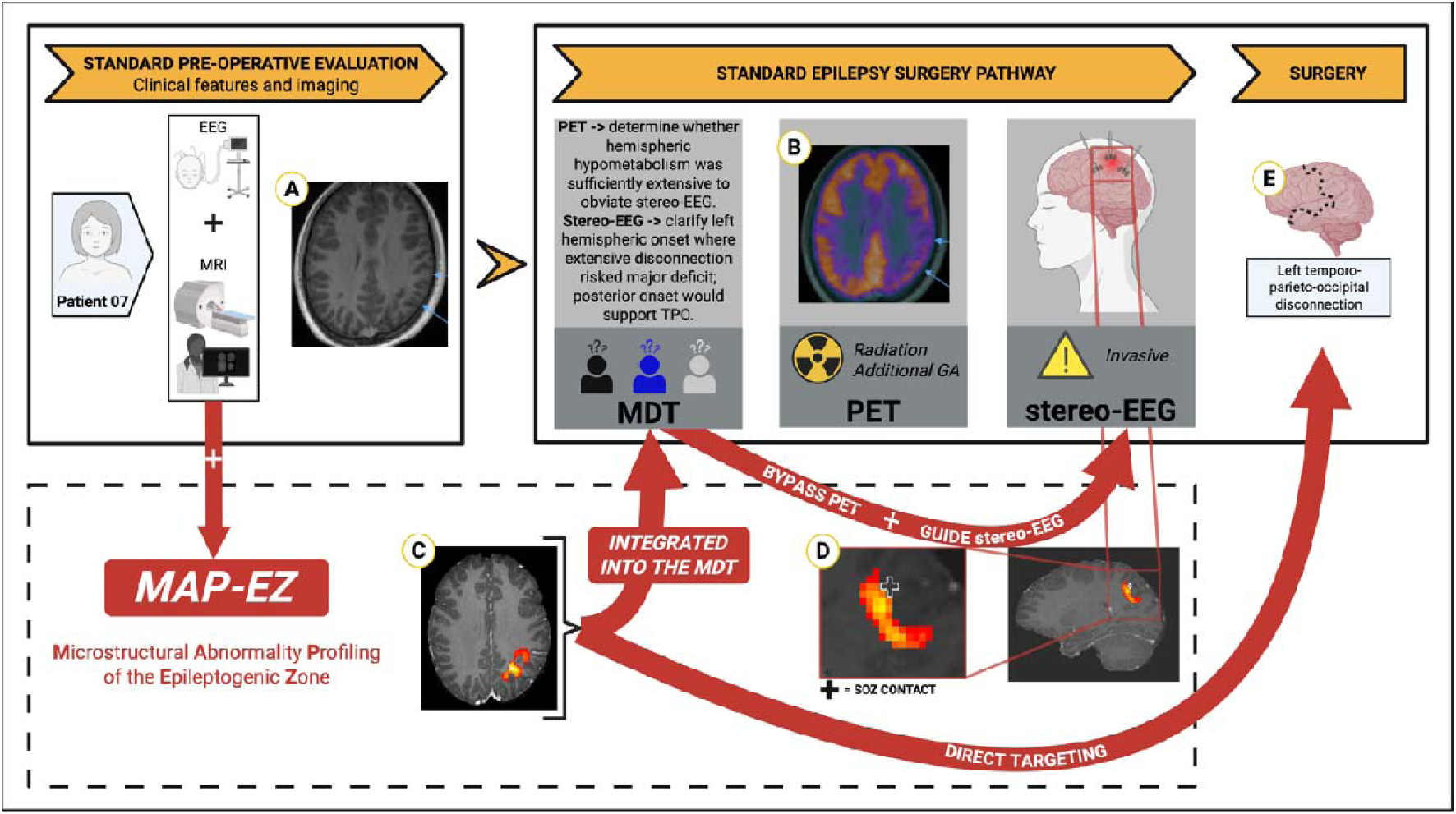
Overview of stages in the patient pathway where microstructural abnormality profiling of the epileptogenic zone (*MAP-EZ*) supports conventional investigations. In patient 07, MAP-EZ mapping could potentially substitute PET and guide stereo-EEG. In this case, a focal increase in mean diffusivity was concordant with PET hypometabolism and could inform surgical planning (TPO disconnection). **(A-E)** relate to patient 07: with Beckwith–Wiedemann syndrome; seizure onset in infancy (right focal sensorimotor with generalization). EEG showed interictal spikes maximal over the left posterior temporal and frontal regions; semiology was poorly localizing. Neuropsychology: high-average verbal reasoning with intact memory/language. fMRI showed right frontal expressive activation and predominantly left temporal comprehension. Visual fields normal. **(A)** MRI: diffuse left hemispheric atrophy/scarring, maximal occipital. **(B)** PET rationale: determine whether hemispheric hypometabolism was sufficiently extensive to obviate stereo-EEG. PET: left anterior parietal hypometabolism extending to postcentral, supramarginal, lateral sulcus, and angular gyrus. **(C)** MAP-EZ: focal subcortical MD hotspot (overlaid on the T1 scan) concordant with PET hypometabolism. **(D)** stereo-EEG rationale: clarify onset within a presumed left hemispheric network where extensive disconnection risked major deficit; posterior onset would support TPO. stereo-EEG: variable posterior onset (superior parietal/inferior temporal) with widespread excitability and non-habitual stimulation responses, suggesting a broad SOZ. **(E)** Surgery: TPO disconnection sparing anterior temporal/mesial structures and language cortex. The subsequent histological diagnosis was FCDIIb and the patient was subsequently SF.

### Diffusion MRI abnormalities localize the EZ and are concordant with PET hypometabolism and stereo-EEG findings

We found that MD is more informative than FA for detecting clinically relevant diffusion abnormalities in focal epilepsy, consistent with prior work.^12^ Although increases in MD have been reported to be more extensive than decreases in FA,^7^ neither metric should be interpreted in isolation. For example, in our cohort of 110 patients who underwent resective surgery, 28 patients had no amount of their dominant MD abnormality resected (selected based on the hypothesized SOZ lobe). Of these, 12 had partial resection of an additional large abnormal cluster derived from the positive/negative MD/FA z-maps. One such patient is shown in Supplementary Figure 14 – a patient who underwent left frontal lesionectomy for an FCD IIb, subsequently achieving seizure freedom. The abnormality was manifested as a decrease in MD (rather than an expected increase). One potential explanation is that peri-ictal changes may have influenced the dMRI signal, as MD is sensitive to ictal activity and shows a biphasic response, with an initial decrease attributed to excitotoxic cellular swelling followed by a later increase.^29,30^

We therefore visually reviewed diffusion maps across all modalities in the remaining 16 patients to identify additional, informative abnormalities. Lateralizing abnormalities were present in 9 patients and provided additional information in 2 out of 3 of these who were NSF after surgery. One patient with persistent seizures after left anterior temporal lobectomy and amygdalohippocampectomy was later considered for temporo-occipito-parietal disconnection; preoperative MD and FA maps showed a well-lateralized parieto-occipital abnormality that may have supported the initial localization and subsequent consideration for re-operation (Supplementary Fig. 13). Our findings differ from earlier dMRI studies in patients with TLE, where interictal apparent diffusion coefficient did not offer lateralizing information for those with non-lateralizing conventional MRI.^31^ That study concluded that interictal DWI did not aid in deciding on temporal lobectomy, as it provided no extra lateralizing information beyond standard epilepsy imaging. This may, in part, relate to the focus of their analysis on apparent diffusion coefficient values in hippocampal regions of interest, whereas our approach assessed diffusion abnormalities voxel-wise across the brain and incorporated both MD and FA. In our study, diffusion mapping provided additive information to the existing MRI and PET findings. For example, a diffusion abnormality was found in the region of the posterior hippocampus, adjacent to the posterior margin of the resection in patient 26, whose histological diagnosis was HS, and they were subsequently NSF (Supplementary Fig. 12). These cases highlight the value of manual review of individual diffusion maps and the need to better characterize how the direction, magnitude and significance of abnormalities differ across pathologies, including FCDs, where reported dMRI properties vary.^32,33^

In MRI lesion-negative patients, diffusion abnormalities were spatially concordant with PET hypometabolism and stereo-EEG findings. The diffusion abnormalities were mostly in the WM adjacent to or underlying the cortical PET hypometabolism (e.g. patients 06 and 07). This raises the possibility that, particularly in FCDs, diffusion abnormalities may reflect adjacent or underlying white matter involvement rather than direct overlap with the strictly cortical component of the malformation. Studies combining FDG-PET with dMRI show that regions of PET hypometabolism are associated with microstructural diffusion abnormalities, particularly in adjacent or connected white-matter pathways.^34–36^ FDG-PET also overlaps with stereo-EEG-defined epileptogenic regions, but hypometabolic regions can be broader than the EZ and may include propagation zones or wider network effects.^37–41^ Connectivity between dominant normative dMRI abnormalities and regions of PET hypometabolism requires further characterization. Integrating arterial spin labelling^42^ and intracranial EEG^43^ with imaging features can further increase localization yield. Such multimodal approaches may define a proximal-to-distal epileptogenic network, extending from a cortical focus to propagating WM, and could inform combined resection and disconnection strategies.

### Resecting diffusion MRI abnormalities predicts seizure freedom

Resecting approximately 36% of the dominant MD abnormality provided the best statistical separation between SF and NSF patients in ROC analysis, although this threshold showed wide variability on bootstrap resampling and should not be interpreted as a strict clinical cutoff. Instead, the findings indicate a general trend whereby greater resection of diffusion-defined abnormal tissue is associated with higher odds of seizure freedom. This is broadly consistent with recent work reporting that removal of even a small proportion of the largest diffusion cluster was associated with a good outcome, but that very extensive resections (≥70%) did not confer additional benefit over more modest resections (<30%).^12^ Together, these observations suggest a positive but non-linear relationship between extent of diffusion abnormality resection and seizure outcome rather than a single optimal percentage. One possible interpretation is that this non-linear pattern reflects a network-level phenomenon, whereby resection of a sufficient proportion of abnormal tissue disrupts the epileptogenic network beyond a critical threshold. Beyond this point, additional resection may confer diminishing benefit, either because the key network components have already been removed or because the remaining abnormal tissue is no longer sufficiently connected to sustain seizures.

### Seizure onset contacts have more abnormal diffusion MRI metrics than uninvolved gray matter in patients undergoing stereo-EEG

In the stereo-EEG cohort, MD z-scores were higher, and FA z-scores lower in seizure onset contacts than in remaining GM contacts, with interictal contacts intermediate. Moreover, SF patients had more abnormal MD and FA values at seizure onset and GM contacts compared to NSF patients however statistical comparison was limited due to the subset of patients included in this analysis (Supplementary Fig. 6). This supports the notion that seizure-generating tissue demonstrates greater microstructural disruption.^9,44,45^ However, a growing body of evidence implicates the WM in seizure propagation and network-level epileptogenesis.^46–48^ In our study a greater proportion of the diffusion abnormalities were in the WM compared to GM and were often focal and underlying the resected and/or ablated region, e.g. patients 02, 09 and 18 (Supplementary Figs. 9,11). This should be interpreted in the context of the conventional surgical paradigm, in which epilepsy surgery is primarily directed towards removing or disconnecting epileptogenic GM. GM diffusion abnormalities may therefore be more directly relevant for identifying a resectable epileptogenic focus, whereas WM abnormalities may reflect altered connectivity, propagation pathways, or broader network disruption rather than a discrete surgical target.

It has been suggested that stereo-EEG sampling of WM is like ‘tapping’ a telephone wire and could identify the ends of a WM tract that show aberrant activity even if the overlying GM region is not sampled.^48^ Thus, dMRI mapping has the potential to help guide the placement of stereo-EEG electrodes, e.g. patient 09 (Supplementary Fig. 9), where electrodes were placed in the vicinity of but not within or adjacent to the diffusion abnormality in the right frontal region. In epilepsy surgery meetings, where the aim is often to identify a resectable GM lesion or focus, the proportion of GM abnormality resected may be the more clinically relevant measure. By contrast, WM diffusion abnormalities may be particularly useful for understanding network architecture and connectivity in more complex developmental pathologies, such as tuberous sclerosis complex or GM heterotopia, where epileptogenicity may depend on distributed cortico-subcortical networks rather than a single focal cortical abnormality.

### Patients without an identified SOZ exhibit more widespread, contralateral and bilateral diffusion MRI abnormalities

In the stereo-EEG cohort, SOZ-not-identified patients exhibited higher MD abnormalities contralateral to the presumed SOZ compared to patients with an identified SOZ, suggesting a more distributed or less localizable disease process. In contrast, patients in whom the SOZ was identified had higher abnormalities ipsilaterally. These findings allude to the potential benefit of dMRI mapping in identifying contralateral and global diffusion abnormalities that could indicate patients in whom stereo-EEG is unlikely to be successful in determining the SOZ. These patterns support the argument that more widespread WM changes relate to more extended seizure networks, which are associated with poorer outcomes. Moreover, this supports recent findings that patients with poor outcomes had higher functional connectivity between the putative SOZ and more distant WM recordings compared to patients with good outcomes.^48^

### Limitations

Although this was a retrospective single-center study, the multi-shell dMRI protocol has formed part of the routine neuroimaging protocol at GOSH since 2015, thereby minimizing potential selection bias. Moreover, all scans were acquired on the same 3T scanner using an identical protocol, reducing technical variance. Three patients had minor acquisition deviations although prior work suggests limited impact on diffusion metrics.^49^ Given that our study utilized high-quality, multi-shell dMRI, future work should assess whether the present findings can be replicated using only the b=1000 s/mm² shell, as this would reduce acquisition time and improve feasibility for centers that acquire single-shell DTI only.

TFCE parameters were used to enhance patient-level maps weighting voxels by their intensity and spatial extent. This approach does not use group-level permutation-based inference to derive family-wise error corrected *P*-values, which can be insensitive to patient-specific effects and may obscure clinically meaningful individual variability^50^, instead focusing on dominant patient-specific abnormalities. In defining the dominant abnormality cluster, the resection lobe was used as a surrogate for the preoperative hypothesized SOZ lobe. This approach assumes that the lobe targeted for resection reflects the multidisciplinary team’s presurgical hypothesis regarding the location of the epileptogenic zone and seizure onset zone. Alternative approaches were explored, including selecting the largest cluster irrespective of location and assessing the proportion resected; however, the resection-lobe approach was chosen because it provided a semi-automated method for identifying the clinically relevant cluster while avoiding manual selection, which could have been biased by knowledge of the resection mask.

Concordance with PET hypometabolism and stereo-EEG findings was assessed descriptively at a regional level and should therefore be interpreted as supportive evidence requiring prospective validation. The median post-surgical follow-up was 21.4 months (range 11.1– 100.9) so it is possible that some children classified as SF may experience a late recurrence.

## Conclusion

This study supports patient-level dMRI for EZ mapping in children with drug-resistant epilepsy. dMRI identified abnormalities that (i) correspond with invasive electrophysiological findings, (ii) differentiate between patients in whom stereo-EEG identified an SOZ and those in whom no SOZ was identified, (iii) show an association between resection of these abnormalities and subsequent seizure freedom, and (iv) localize to the seizure onset zone even in patients without visible MRI lesions. Together, these findings highlight the potential value of normative dMRI as a complementary component of the pre-operative epilepsy surgery pathway with potential to refine presurgical localization, guide stereo-EEG implantation, support surgical candidate selection, and improve outcome prediction.

## Data availability

Derived data supporting the findings of this study are available from the corresponding author on request. The scripts to process the data will be made available via GitHub (https://github.com/veljanoskid/EZ-MAP-Epileptogenic-Zone-Microstructural-Abnormality-Profiling)

## Acknowledgements

We thank Jonathan Horsley, Peter Taylor, Gerard Hall and Karoline Leiberg of the Computational Neurology, Neuroscience and Psychiatry Lab at Newcastle University for helpful discussions on normative modelling and diffusion MRI preprocessing. A large language model was used to support script development and decoding.

## Funding

This work is supported by the NIHR GOSH Biomedical Research Centre. DV is funded by the Great Ormond Street Hospital Children’s Charity Lewis Spitz Surgeon-Scientist PhD Programme. AC and RJP are supported by NIHR Academic Clinical Lectureships. RJP is supported by the GOSH Children’s Charity.

## Competing interests

The authors report no competing interests.

## Author contribution statement

D.V., A.C., C.A.C., M.M.T., T.B., and R.J.P contributed to the conception and design of the study; all authors contributed to acquisition or analysis of data, D.V., A.C., C.A.C., F.D., K.M., M.M.T., T.B., and R.J.P contributed to drafting the text or preparing the figures.

## Abbreviations

dMRI: Diffusion magnetic resonance imaging
DRE: Drug-resistant epilepsy
EZ: Epileptogenic zone
FA: Fractional anisotropy
FCD: Focal cortical dysplasia
FDG-PET: Fluorodeoxyglucose positron emission tomography
FLAIR: Fluid-attenuated inversion recovery
GM: Gray matter
HS: Hippocampal sclerosis
MAP-EZ: Microstructural Abnormality Profiling of the Epileptogenic Zone
MD: Mean diffusivity
NSF: Not seizure-free
*P_perm_*: Permutation *P*-value
SF: Seizure-free
Stereo-EEG: Stereoelectroencephalography
SOZ: Seizure onset zone
TFCE: Threshold-free cluster enhancement
TLE: Temporal lobe epilepsy
WM: White matter

## Notes

### Competing Interest Statement

The authors have declared no competing interest.

### Author Declarations

This retrospective observational study was approved by the Research Governance Office of Great Ormond Street Hospital (ID: 23NP01). The dataset used in the study was de-identified prior to analysis.

## References

1. Eriksson MH, Whitaker KJ, Booth J, et al. Pediatric epilepsy surgery from 2000 to 2018: Changes in referral and surgical volumes, patient characteristics, genetic testing, and postsurgical outcomes. Epilepsia. 2023;64(9):2260–2273. doi:10.1111/epi.17670

2. Widjaja E, Jain P, Demoe L, Guttmann A, Tomlinson G, Sander B. Seizure outcome of pediatric epilepsy surgery. Neurology. 2020;94(7):311–321. doi:10.1212/WNL.0000000000008966

3. UK Children’s Epilepsy Surgery Collaboration. The UK experience of stereoelectroencephalography in children: An analysis of factors predicting the identification of a seizure-onset zone and subsequent seizure freedom. Epilepsia. 2021;62(8):1883–1896. doi:10.1111/epi.16954

4. Basser PJ, Mattiello J, LeBihan D. MR diffusion tensor spectroscopy and imaging. Biophys J. 1994;66(1):259–267. doi:10.1016/S0006-3495(94)80775-1

5. Wieshmann UC, Clark CA, Symms MR, Franconi F, Barker GJ, Shorvon SD. Reduced anisotropy of water diffusion in structural cerebral abnormalities demonstrated with diffusion tensor imaging. Magnetic Resonance Imaging. 1999;17(9):1269–1274. doi:10.1016/S0730-725X(99)00082-X

6. Wieshmann UC, Clark CA, Symms MR, Barker GJ, Birnie KD, Shorvon SD. Water diffusion in the human hippocampus in epilepsy. Magnetic Resonance Imaging. 1999;17(1):29–36. doi:10.1016/S0730-725X(98)00153-2

7. Eriksson SH, Rugg-Gunn FJ, Symms MR, Barker GJ, Duncan JS. Diffusion tensor imaging in patients with epilepsy and malformations of cortical development. Brain. 2001;124(3):617–626. doi:10.1093/brain/124.3.617

8. Le Bihan D, Mangin JF, Poupon C, et al. Diffusion tensor imaging: concepts and applications. J Magn Reson Imaging. 2001;13(4):534–546. doi:10.1002/jmri.1076

9. Winston GP, Vos SB, Caldairou B, et al. Microstructural imaging in temporal lobe epilepsy: Diffusion imaging changes relate to reduced neurite density. Neuroimage Clin. 2020;26:102231. doi:10.1016/j.nicl.2020.102231

10. Thivard L, Adam C, Hasboun D, et al. Interictal diffusion MRI in partial epilepsies explored with intracerebral electrodes. Brain. 2006;129(2):375 EP–385. doi:10.1093/brain/awh709

11. Thivard L, Bouilleret V, Chassoux F, et al. Diffusion tensor imaging can localize the epileptogenic zone in nonlesional extra-temporal refractory epilepsies when [18F]FDG-PET is not contributive. Epilepsy Research. 2011;97(1-2):170–182. doi:10.1016/j.eplepsyres.2011.08.005

12. Horsley J, Hall G, Simpson C, et al. Seizure freedom after surgical resection of diffusion-weighted magnetic resonance imaging abnormalities. Epilepsia. 2025;66(9):3480–3490. doi:10.1111/epi.18490

13. Lebel C, Gee M, Camicioli R, Wieler M, Martin W, Beaulieu C. Diffusion tensor imaging of white matter tract evolution over the lifespan. NeuroImage. 2012;60(1). doi:10.1016/j.neuroimage.2011.11.094

14. Reynolds JE, Grohs MN, Dewey D, Lebel C. Global and regional white matter development in early childhood. NeuroImage. 2019;196:49–58. doi:10.1016/j.neuroimage.2019.04.004

15. Chari A, Seunarine KK, He X, et al. Drug-resistant focal epilepsy in children is associated with increased modal controllability of the whole brain and epileptogenic regions. Commun Biol. 2022;5(1):394. doi:10.1038/s42003-022-03342-8

16. Stotesbury H, Kirkham FJ, Kölbel M, et al. White matter integrity and processing speed in sickle cell anemia. Neurology. 2018;90(23):e2042–e2050. doi:10.1212/WNL.0000000000005644

17. Barona M, Brown M, Clark C, Frangou S, White T, Micali N. White matter alterations in anorexia nervosa: Evidence from a voxel-based meta-analysis. Neurosci Biobehav Rev. 2019;100:285–295. doi:10.1016/j.neubiorev.2019.03.002

18. Cooper HE, Kaden E, Halliday LF, et al. White matter microstructural abnormalities in children with severe congenital hypothyroidism. Neuroimage Clin. 2019;24:101980. doi:10.1016/j.nicl.2019.101980

19. Cruces RR, Royer J, Herholz P, et al. Micapipe: A pipeline for multimodal neuroimaging and connectome analysis. NeuroImage. 2022;263:119612. doi:10.1016/j.neuroimage.2022.119612

20. Rutherford S, Marquand AF. Normative Modeling with the Predictive Clinical Neuroscience Toolkit (PCNtoolkit). In: Whelan R, Lemaître H, eds. Methods for Analyzing Large Neuroimaging Datasets. Springer US; 2025:329–364. doi:10.1007/978-1-0716-4260-3_14

21. Smith SM, Nichols TE. Threshold-free cluster enhancement: Addressing problems of smoothing, threshold dependence and localisation in cluster inference. NeuroImage. 2009;44(1):83–98. doi:10.1016/j.neuroimage.2008.03.061

22. Rugg-Gunn FJ. Diffusion tensor imaging of cryptogenic and acquired partial epilepsies. Brain. 2001;124(3):627–636. doi:10.1093/brain/124.3.627

23. Rugg-Gunn FJ, Eriksson SH, Symms MR, et al. Diffusion tensor imaging in refractory epilepsy. The Lancet. 2002;359(9319):1748–1751. doi:10.1016/S0140-6736(02)08615-4

24. Guye M, Ranjeva JP, Bartolomei F, et al. What is the significance of interictal water diffusion changes in frontal lobe epilepsies? NeuroImage. 2007;35(1):28–37. doi:10.1016/j.neuroimage.2006.11.049

25. Narizzano M, Arnulfo G, Ricci S, et al. SEEG assistant: a 3DSlicer extension to support epilepsy surgery. BMC Bioinformatics. 2017;18(1):124. doi:10.1186/s12859-017-1545-8

26. Modat M, Cash DM, Daga P, Winston GP, Duncan JS, Ourselin S. Global image registration using a symmetric block-matching approach. J Med Imaging (Bellingham). 2014;1(2):024003. doi:10.1117/1.JMI.1.2.024003

27. Khan M, Chari A, Seunarine K, et al. Proportion of resected seizure onset zone contacts in pediatric stereo-EEG-guided resective surgery does not correlate with outcome. Clinical Neurophysiology. 2022;138:18–24. doi:10.1016/j.clinph.2022.03.012

28. Ho J, Tumkaya T, Aryal S, Choi H, Claridge-Chang A. Moving beyond P values: data analysis with estimation graphics. Nat Methods. 2019;16(7):565–566. doi:10.1038/s41592-019-0470-3

29. Chu K, Kang DW, Kim JY, Chang KH, Lee SK. Diffusion-Weighted Magnetic Resonance Imaging in Nonconvulsive Status Epilepticus. Arch Neurol. 2001;58(6):993–998. doi:10.1001/archneur.58.6.993

30. Diehl B, Symms MR, Boulby PA, et al. Postictal diffusion tensor imaging. Epilepsy Research. 2005;65(3):137–146. doi:10.1016/j.eplepsyres.2005.05.007

31. Wehner T, Lapresto E, Tkach J, et al. The value of interictal diffusion-weighted imaging in lateralizing temporal lobe epilepsy. Neurology. 2007;68(2):122–127. doi:10.1212/01.wnl.0000250337.40309.3d

32. Lorio S, Adler S, Gunny R, et al. MRI profiling of focal cortical dysplasia using multi compartment diffusion models. Epilepsia. 2020;61(3):433–444. doi:10.1111/epi.16451

33. Gennari AG, Cserpan D, Stefanos-Yakoub I, Kottke R, O’Gorman Tuura R, Ramantani G. Diffusion tensor imaging discriminates focal cortical dysplasia from normal brain parenchyma and differentiates between focal cortical dysplasia types. Insights Imaging. 2023;14(1):36. doi:10.1186/s13244-023-01368-y

34. Lippe S, Poupon C, Cachia A, et al. White matter abnormalities revealed by DTI correlate with interictal grey matter FDG-PET metabolism in focal childhood epilepsies. Epileptic Disorders. 2012;14(4):404 EP–413. doi:10.1684/epd.2012.0547

35. Poirier SE, Kwan BYM, Jurkiewicz MT, et al. 18F-FDG PET-guided diffusion tractography reveals white matter abnormalities around the epileptic focus in medically refractory epilepsy: implications for epilepsy surgical evaluation. European J Hybrid Imaging. 2020;4(1):10. doi:10.1186/s41824-020-00079-7

36. Tatekawa H, Uetani H, Hagiwara A, et al. Association of hypometabolic extension of 18F-FDG PET with diffusion tensor imaging indices in mesial temporal lobe epilepsy with hippocampal sclerosis. Seizure. 2021;88:130–137. doi:10.1016/j.seizure.2021.04.007

37. Lucignani G, Tassi L, Fazio F, et al. Double-blind stereo-EEG and FDG PET study in severe partial epilepsies: are the electric and metabolic findings related? Eur J Nucl Med. 1996;23(11):1498–1507. doi:10.1007/BF01254475

38. Lamarche F, Job AS, Deman P, et al. Correlation of FDG-PET hypometabolism and SEEG epileptogenicity mapping in patients with drug-resistant focal epilepsy. Epilepsia. 2016;57(12):2045–2055. doi:10.1111/epi.13592

39. Popescu CE, Mai R, Sara R, et al. The Role of FDG-PET in Patients with Epilepsy Related to Periventricular Nodular Heterotopias: Diagnostic Features and Long-Term Outcome. J Neuroimaging. 2019;29(4):512–520. doi:10.1111/jon.12620

40. Lagarde S, Boucekine M, McGonigal A, et al. Relationship between PET metabolism and SEEG epileptogenicity in focal lesional epilepsy. Eur J Nucl Med Mol Imaging. 2020;47(13):3130–3142. doi:10.1007/s00259-020-04791-1

41. Zhang L, Zhou H, Tang Y, et al. Identifying the epileptogenic zone by 18F-FDG PET/MRI in drug-resistant epilepsy with focal cortical dysplasia type IIIa. Clinical Neurophysiology. 2026;181:2111412. doi:10.1016/j.clinph.2025.2111412

42. Gennari AG, Cserpan D, Kottke R, et al. Arterial spin labeling performs comparably to 2-[18F]fluoro-2-deoxy-D-glucose positron emission tomography for presurgical evaluation in pediatric lesional epilepsy. Epilepsia. Published online March 14, 2026. doi:10.1002/epi.70199

43. Kozma C, Horsley J, Hall G, et al. Multimodal integration of magnetic resonance imaging and intracranial electroencephalographic abnormalities in temporal lobe epilepsy surgery. Epilepsia. 2026;67(3):1181–1192. doi:10.1111/epi.70042

44. Govindan RM, Asano E, Juhasz C, Jeong J won, Chugani HT. Surface based laminar analysis of diffusion abnormalities in cortical and white matter layers in neocortical epilepsy. Epilepsia. 2013;54(4):667–677. doi:10.1111/epi.12129

45. Sahlas E, Avigdor T, Ngo A, et al. Alterations in Cortical Microstructure, Morphology, and Intrinsic Local Function in Spiking Tissue in Patients With Focal Epilepsy. Neurology. 2025;104(12):e213733. doi:10.1212/WNL.0000000000213733

46. Otte WM, van Eijsden P, Sander JW, Duncan JS, Dijkhuizen RM, Braun KPJ. A meta-analysis of white matter changes in temporal lobe epilepsy as studied with diffusion tensor imaging. Epilepsia. 2012;53(4):659–667. doi:10.1111/j.1528-1167.2012.03426.x

47. Pruckner P, Mito R, Vaughan DN, et al. Surgical white matter disruption leads to downstream atrophy in the non-resected human brain. Brain. 2026;149(2):548–562. doi:10.1093/brain/awaf344

48. Revell AY, Jaskir M, Lucas A, et al. White matter signals reflect information transmission between brain regions during seizures. Brain. 2026;149(1):77–89. doi:10.1093/brain/awaf444

49. Grech-Sollars M, Hales PW, Miyazaki K, et al. Multi-centre reproducibility of diffusion MRI parameters for clinical sequences in the brain. NMR in Biomedicine. 2015;28(4):468–485. doi:10.1002/nbm.3269

50. Mito R, Pedersen M, Pardoe H, et al. Exploring individual fixel-based white matter abnormalities in epilepsy. Brain Commun. 2023;6(1):fcad352. doi:10.1093/braincomms/fcad352

