## Supplementary file for "Diffusion MRI abnormalities localise the epileptogenic zone in paediatric drug-resistant epilepsy"

Supplementary Materials

Please see supplemental materials below.

### Supplementary Methods:

#### 1. MRI processing

MRI data were acquired on a single Siemens Magnetom Prisma 3T scanner at GOSH using either a 20- or 64-channel head coil, and a multi-shell diffusion protocol (b=1000 and 2200s/mm^2^) with consistent acquisition parameters across all control and patient cohorts. A spin-echo single-shot 2D EPI acquisition and multiband (factor 2) sequence with 60 non-collinear diffusion directions per shell were used. Acquisition parameters were highly consistent across the cohorts, with minor deviations in three patients (slightly longer repetition times in two patients, marginal differences in slice gap in two patients, and a slightly larger in-plane voxel size (2.09×2.09mm) in one patient due to field-of-view and matrix differences). The typical dMRI spatial resolution was 2mm in-plane with a 0.2mm gap across 66 slices. TR=3050ms, TE=60ms, field of view=220mm×220mm, matrix size=110×110, in-plane voxel resolution=2.0mmx2.0mm, GRAPPA factor 2, phase-encoding partial Fourier=6/8. An additional b=0 scan with identical readout but 180° anterior-posterior phase-encode reversal was acquired for susceptibility-related artefact correction.

DICOM files were converted to BIDS format using BIDSCoin (version 4.6.2). Structural and dMRI preprocessing was performed using the structural (*proc_structural*, *proc_surf*, *post_structural*) and diffusion (*proc_dwi*) modules of micapipe (version 0.2.3) with standard settings.^29^ A population-average FA template was built from the control cohort with the *population_template* tool^61^ from MRtrix using centre-of-mass alignment, a voxel size of 2x2x2.2mm, and rigid, affine non-linear registration. This produced a voxelwise mean of all aligned FA images (Fig. 1B). A population-average T1 template was also derived.

For the resective cohort, the preoperative T1 scan was registered to the postoperative T1 scan and the b=0 scan (with brain masks generated using micapipe and/or HD-BET) using *antsRegistrationSyN* (Fig. 1A).^62^ The preoperative T1-weighted scan was used in most patients; in a small minority, a post-contrast T1-weighted scan was substituted where the non-contrast scan was degraded by artefact or lower image quality. The resection masks were manually segmented by authors RJP and DV in native space using ITKSnap (version 4.4.0). *antsApplyTransforms*^62^ with NearestNeighbour interpolation was used to warp the postoperative resection mask into the preoperative T1 space and then into diffusion space. Empty voxels within the resection masks were filled, and Gaussian smoothing followed by re-thresholding at 0.3 was applied to enforce anatomical continuity after registration to template space and to regularise the resection boundary. For all patient cohorts the brain-masked FA maps were registered to the control cohort FA template using *antsRegistrationSyN* (Fig. 1C).^62^ The corresponding transforms were used to warp the MD maps and resection masks to template space.

Non-CSF tissue from each patient’s native five tissue-type image was extracted, and the existing patient-specific FA-to-template transforms were used with nearest-neighbour interpolation to warp the non-CSF mask to template space. Subsequently, cerebellar and brainstem masks were generated from the control cohort T1 template using *deepAtropos,*^63^ and these were also subtracted from each patient’s normative diffusion maps.

#### 2. Group mean maps

For each subgroup, individual MD and FA z-maps were aggregated to produce voxelwise mean and median maps. The mean maps summarise the average spatial pattern of abnormalities and the median maps provide a more outlier-robust representation of the typical patient. The hemispheres of stereo-EEG patients were left-right mirrored so that the ipsilateral hemisphere was consistently displayed on the radiological left side of the resection, SOZ, or primary hypothesis. For all group mean and median maps, voxels were retained only if at least 80% of patients contributed data, otherwise voxels were set to NaN. To assess the prevalence of abnormalities across patients, fraction suprathreshold maps were generated, showing for each voxel the proportion of patients whose z-score exceeded a predefined threshold (MD: positive z-scores; FA: negative z-scores). Between-group difference maps were calculated for the mean, median, and fraction maps, enabling group differences in effect magnitude, robustness, and prevalence to be evaluated in parallel.

#### 3. Statistical methods

To quantify the relationship between the diffusion abnormalities and resection masks, the mean difference of the percentage resected of the dominant diffusion abnormality, derived from MD and FA maps, in SF versus NSF patients was plotted in Gardner-Altman estimation plots as a bootstrap sampling distribution.^64^ The effect sizes and CIs are reported as: effect size (CI width lower bound, upper bound). 5000 bootstrap samples were taken; the confidence interval is bias-corrected and accelerated. The *P*-value reported is the probability of observing the effect size (or greater), assuming the null hypothesis of zero difference is true. For each permutation *P*-value (*P*_perm_), 5000 reshuffles of the SF and NSF groups were performed.

For patients who had at least some of the dominant diffusion abnormality resected, receiver operating characteristic (ROC) curves were estimated and area under the curve (AUC) values were calculated along with nonparametric AUC confidence intervals for MD- and FA-derived abnormalities. The optimal outcome discrimination threshold was defined as the predictor value that maximised the Youden index with the corresponding sensitivity and specificity. Univariable logistic regression models were fit for each metric, scaled per 10 percentage-point increase, using a binomial GLM. HC3 standard errors were used for inference, and odds ratios with Wald 95% confidence intervals were calculated by exponentiating the model coefficients and confidence limits. To quantify uncertainty in the Youden-optimised threshold, nonparametric bootstrap resampling (5,000 iterations, seed=1) was performed separately for each metric. Bootstrap samples were drawn with replacement; resamples with fewer than two outcome classes were excluded. From the retained bootstrap samples, the median Youden-optimised threshold and a 95% confidence interval were reported, along with the median and percentile interval of the bootstrap AUC distribution. Differences in percentage of the dominant diffusion abnormality resected between patients who underwent lobectomy versus lesionectomy were analysed using a two-way ANOVA with a Bonferroni multiple comparison test.

Differences in mean z-scored MD and FA values between GM, interictal and seizure onset contacts in stereo-EEG patients were analysed using paired estimation statistics implemented in Python with the DABEST package. Two repeated measures were performed: 1) paired comparison (GM vs seizure onset contacts) and 2) triplet comparison with a common baseline (GM). For (1), MD values from GM contacts (excluding the seizure onset contacts) were compared with MD values from seizure onset contacts. For (2), MD values from GM contacts (excluding both interictal and seizure onset contacts) were used as a baseline and compared separately against interictal and seizure onset contacts. For all analyses, paired mean differences were estimated using nonparametric bootstrap resampling (5,000 resamples) with bias-corrected and accelerated (BCa) 95% confidence intervals. A fixed random seed was used to ensure reproducibility. Estimation plots were generated for each analysis. For completeness, permutation t-tests (5,000 reshuffles; reported as legacy statistics by DABEST) were also computed and reported.

### Supplementary Figures:

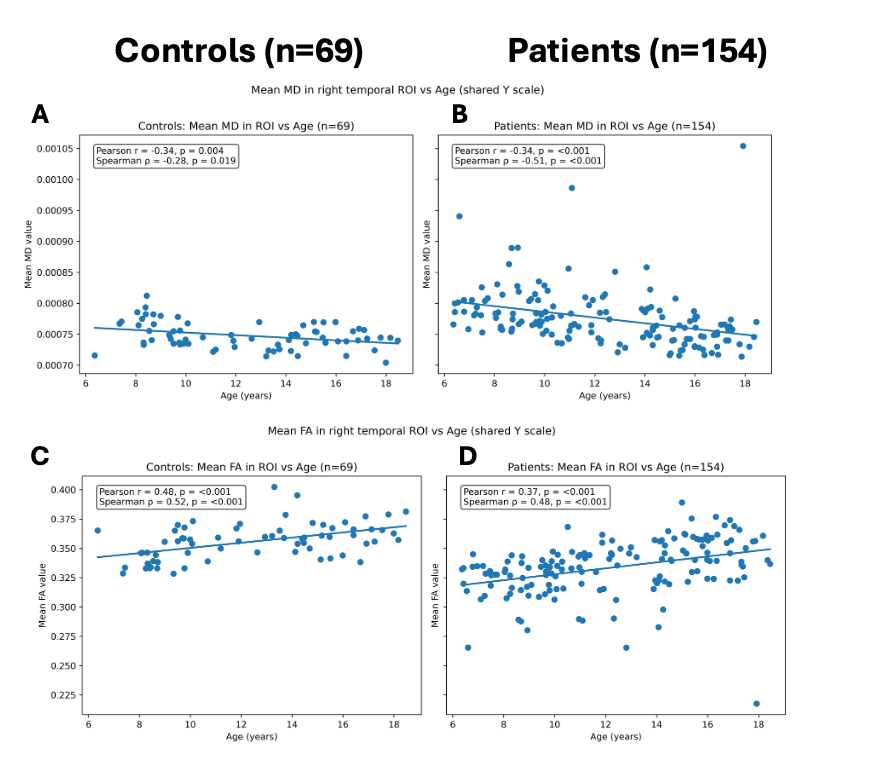

**Supplementary figure 1** Association between mean MD and FA values (in a right temporal region of interest) with age for control and patient cohorts, plotted on a shared scale. **(A)** Mean MD versus age for controls. **(B)** Mean MD versus age for patients. **(C)** Mean FA versus age for controls. **(D)** Mean FA versus age for patients.

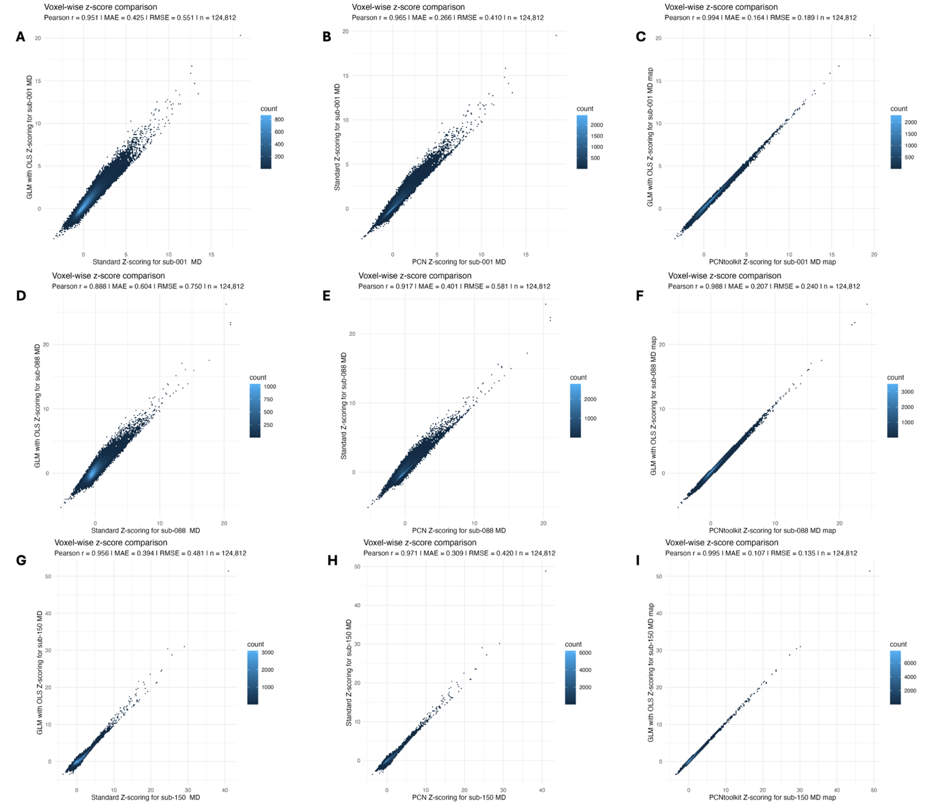

**Supplementary figure 2** Voxelwise comparison of z-scored MD maps generated using standard (raw MD value - mean / standard deviation) z-scoring, GLM (OLS-based) z-scoring (age and sex adjusted), and PCN Toolkit z-scoring (age and sex adjusted) for three patients. **(A)** Voxel-wise comparison between standard (mean–standard deviation) z-scoring and GLM-based z-scoring for patient 33’s mean diffusivity (MD) map. Density bins indicate the number of voxels sharing similar z-score values. Summary statistics (Pearson r, MAE, RMSE) are shown to quantify correspondence. **(B)** Voxel-wise comparison of standard z-scoring and PCN-derived z-scores for patient 33. **(C)** Voxel-wise comparison of z-score maps derived using the PCN toolkit and a GLM with ordinary least squares for patient 33. **(D, E, F)** Comparison plots for patient 16, analogous to **(A, B, C)**. **(G, H, I)** Comparison plots for patient 34.

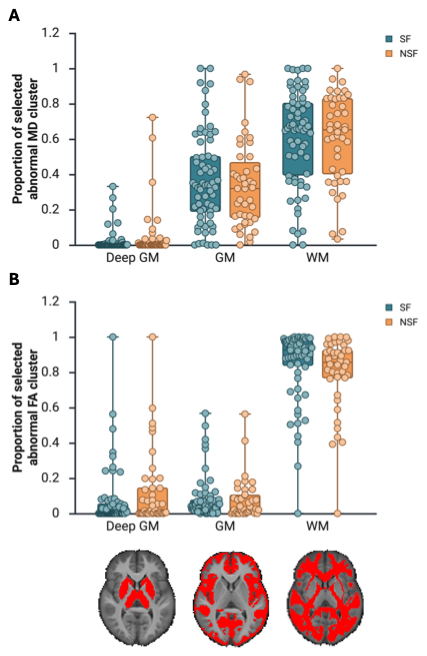

Supplementary figure 3 Proportion of the dominant selected MD (A) and FA (B) abnormality resected stratified by the deep GM, cortical GM, and WM in template space.

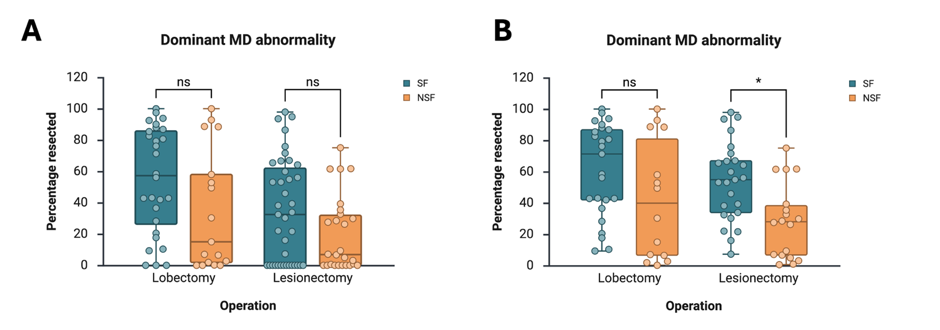

Supplementary figure 4 Percentage resected of the dominant MD diffusion abnormality primarily stratified by type of operation (lobectomy versus lesionectomy) and secondary stratified by seizure freedom. **(A)** Including patients who did not have any amount of the abnormality resected. **(B)** Analysis restricted to patients who had at least some of the abnormality resected.

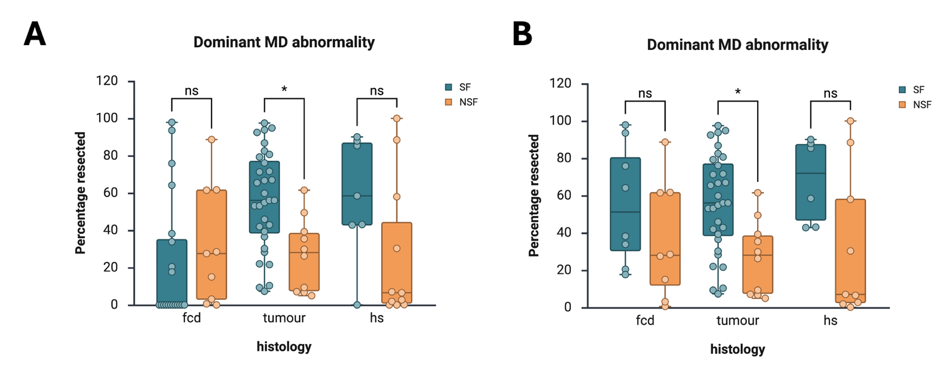

Supplementary figure 5 The relationship between histology (FCD, HS and tumour) and percentage of the dominant MD diffusion abnormality resected, stratified by outcome. (A) Across all patients. (B) Subset of patients with >0% resected.

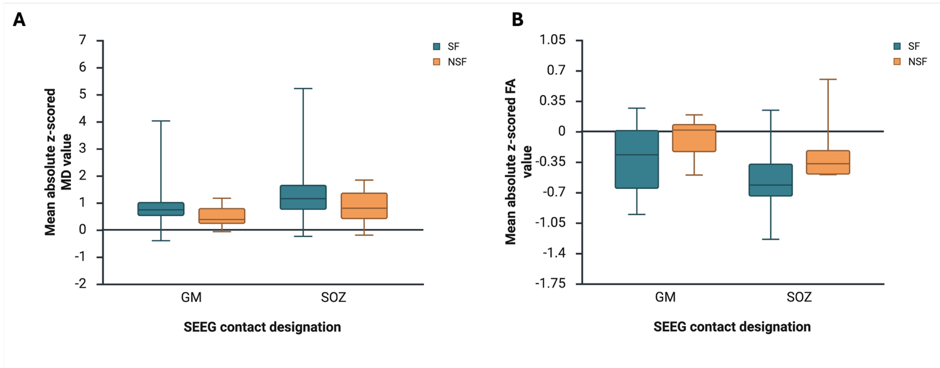

Supplementary figure 6 Mean absolute z-scored values of contacts designated as being in the GM versus seizure onset zone stratified by outcome (A) MD, (B) FA.

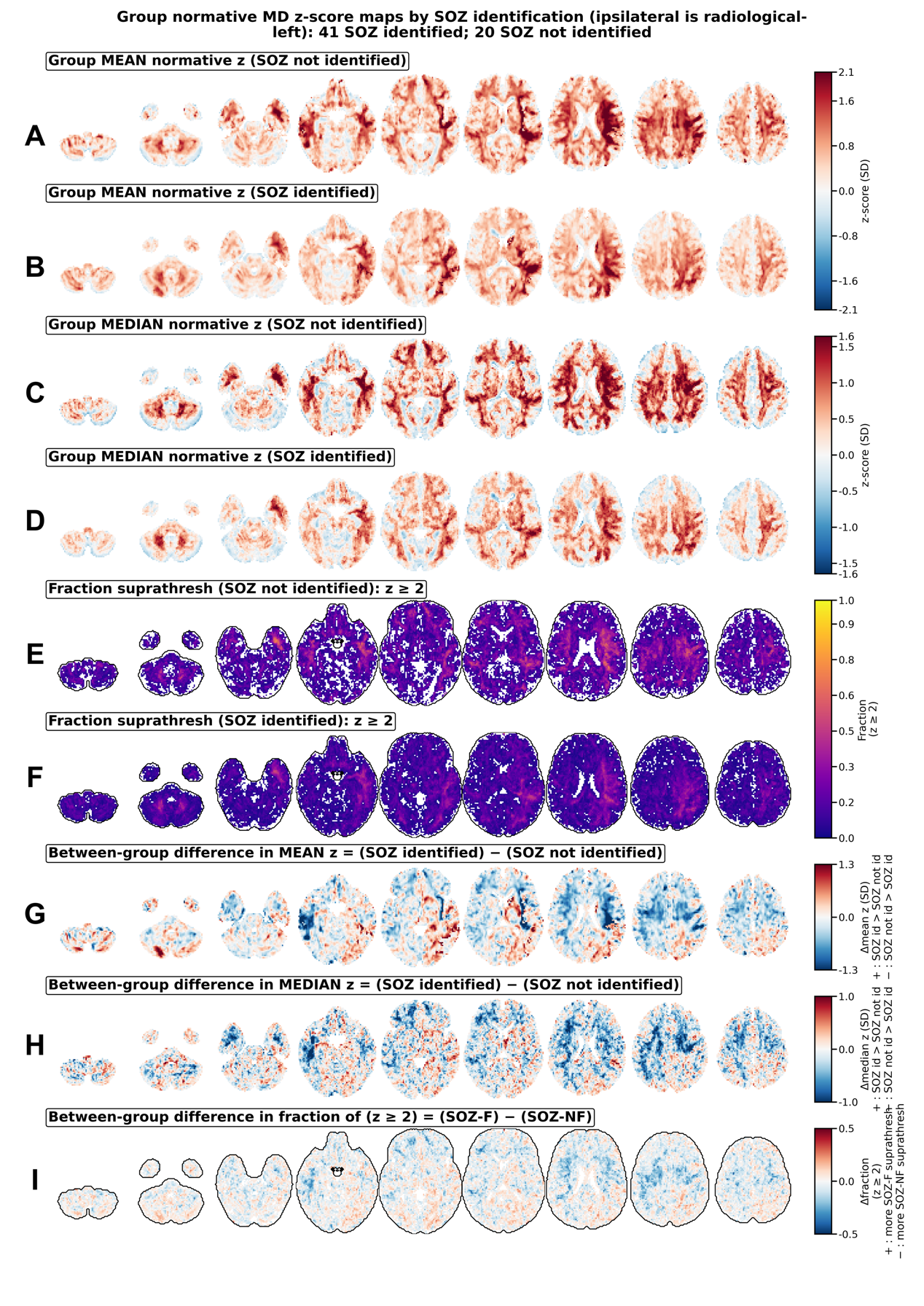

Supplementary figure 7 Group-level normative MD z-score maps are shown for patients with SOZ not identified (n=20) and SOZ identified (n=41), after laterality-based hemispheric flipping (so ipsilateral is radiological left) and the operated/SOZ/primary-hypothesis side appears on the radiological left. Mean **(A-B)** and median **(C-D)** maps are coverage-masked within each group such that voxels are retained only if at least 80% of patients contribute finite data; otherwise, voxels are set to NaN. Fraction suprathreshold maps **(E-F)** show, at each voxel, the proportion of patients exceeding the threshold (z ≥ 2) among patients with finite values at that voxel (voxels with 0 finite subjects are NaN). Between-group mean difference maps **(G)** show Δmean z = (SOZ identified) − (SOZ not identified). To help distinguish whether Δmean effects are driven by a small number of extreme patients versus a broader shift across patients, we also show a Δmedian z map (**H,** less sensitive to outliers) and a Δfraction suprathreshold map (**I,** prevalence of suprathreshold voxels). For visualization, color scales for Δmean and Δmedian maps were set symmetrically using the 99th percentile of the absolute voxel-wise differences, computed separately for mean- and median-based maps. The Δfraction map uses a fixed symmetric scale (-0.5 to +0.5). All maps are displayed in radiological orientation. For plotting only, NaNs are rendered as 0.

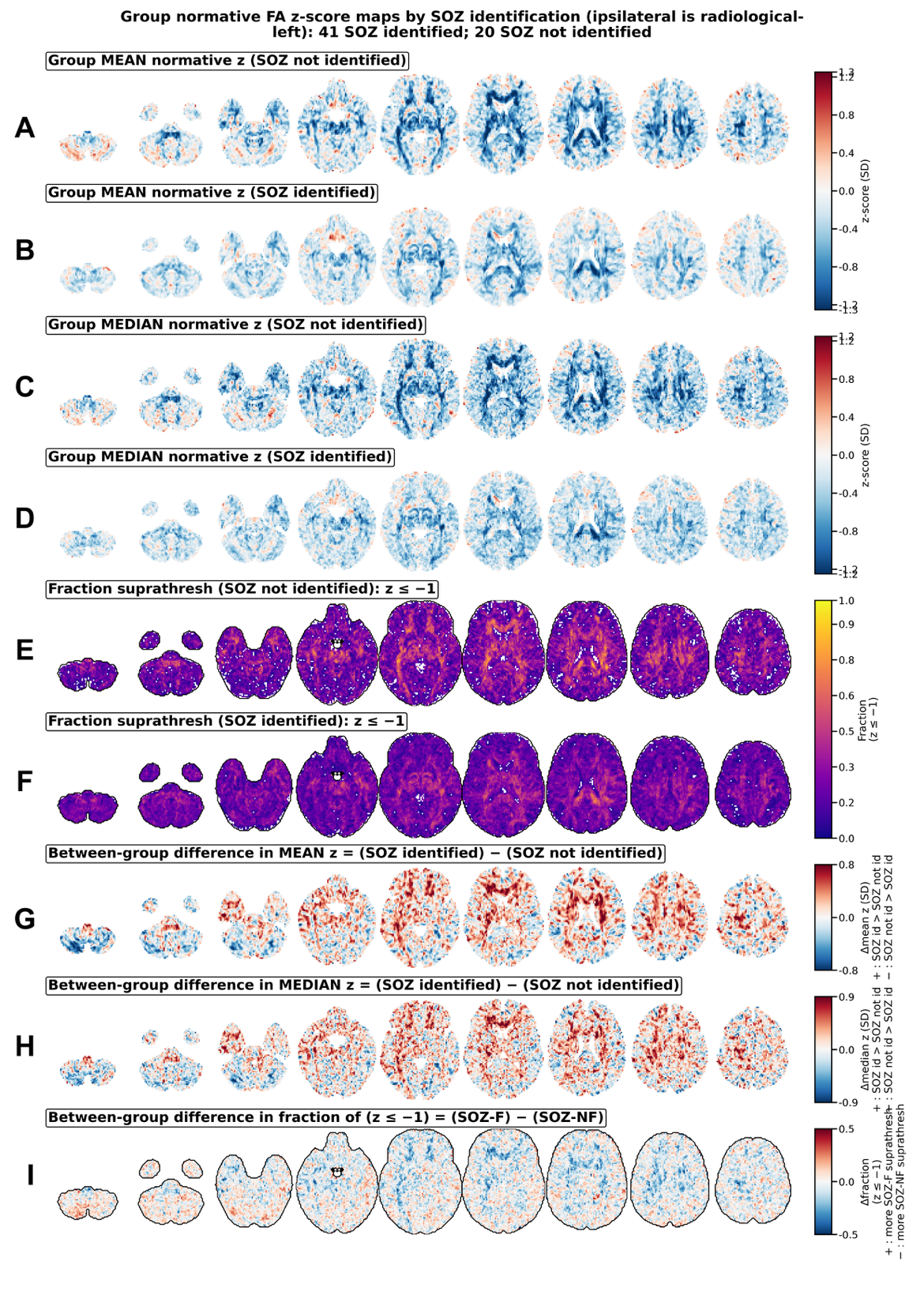

Supplementary figure 8 Group-level normative FA z-score maps are shown for patients with SOZ not identified (n=20) and SOZ identified (n=41), after laterality-based hemispheric flipping (so ipsilateral is radiological left) and the operated/SOZ/primary-hypothesis side appears on the radiological left. Mean **(A-B)** and median **(C-D)** maps are coverage-masked within each group such that voxels are retained only if at least 80% of patients contribute finite data; otherwise, voxels are set to NaN. Fraction suprathreshold maps **(E-F)** show, at each voxel, the proportion of patients exceeding the threshold (z ≤ 1) among patients with finite values at that voxel (voxels with 0 finite patients are NaN). Between-group mean difference maps **(G)** show Δmean z = (SOZ identified) − (SOZ not identified). To help distinguish whether Δmean effects are driven by a small number of extreme patients versus a broader shift across patients, we also show a Δmedian z map (**H,** less sensitive to outliers) and a Δfraction suprathreshold map (**I,** prevalence of suprathreshold voxels). For visualization, color scales for Δmean and Δmedian maps were set symmetrically using the 99th percentile of the absolute voxel-wise differences, computed separately for mean- and median-based maps. The Δfraction map uses a fixed symmetric scale (-0.5 to +0.5). All maps are displayed in radiological orientation. For plotting only, NaNs are rendered as 0.

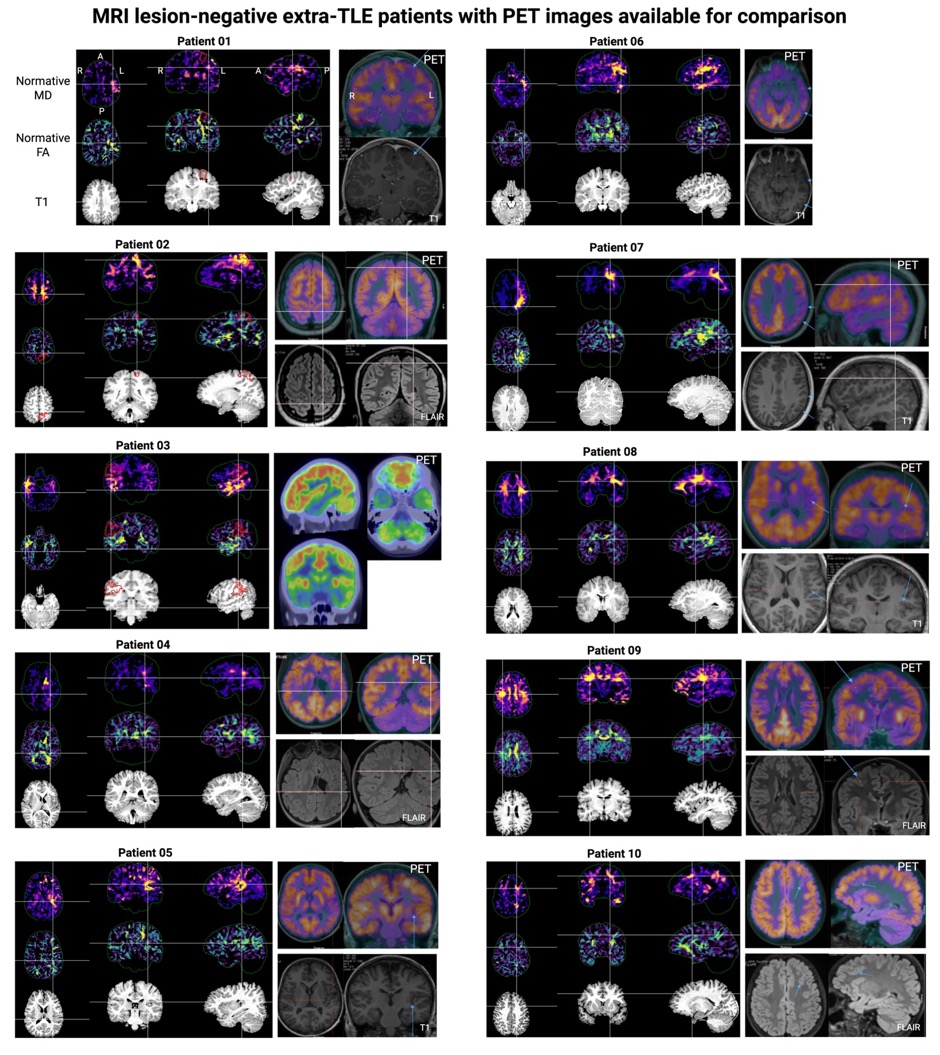

Supplementary figure 9 Per-patient normative diffusion abnormalities in radiological orientation for MRI lesion-negative patients with a confirmed or suspected extra-temporal SOZ with PET images (with or without FLAIR images) for comparison. Left top panel: unthresholded TFCE normative MD map (using positive z-values, reflecting elevated diffusivity relative to controls). Left middle panel: unthresholded TFCE normative FA map (using negative z-values, reflecting reduced anisotropy relative to controls). Left bottom panel: Linearly aligned T1 scan in diffusion template space. Black crosshairs: SOZ-designated stereo-EEG contacts (where applicable). Red outline: resection masks (where applicable). Right panel: PET scan with or without FLAIR/T1 (labelled).

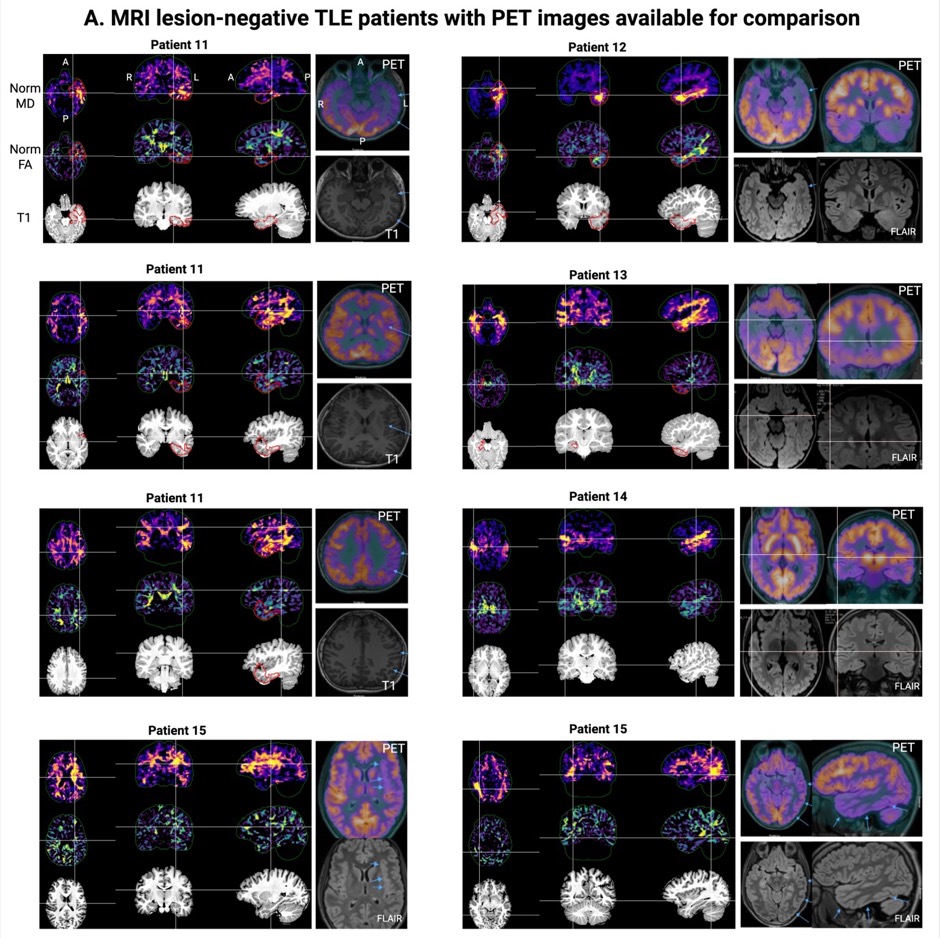

Supplementary figure 10 Per-patient normative diffusion abnormalities in radiological orientation for MRI lesion-negative patients with a confirmed or suspected temporal SOZ with PET images (with or without FLAIR images) for comparison. Left top panel: unthresholded TFCE normative MD map (using positive z-values, reflecting elevated diffusivity relative to controls). Left middle panel: unthresholded TFCE normative FA map (using negative z-values, reflecting reduced anisotropy relative to controls). Left bottom panel: Linearly aligned T1 scan in diffusion template space. Black crosshairs: SOZ-designated stereo-EEG contacts (where applicable). Red outline: resection masks (where applicable). Right panel: PET scan with or without FLAIR/T1 (labelled).

| **Patient** | **Age range; sex** | **Duration of epilepsy (months)** | **MRI diagnosis** | **PET** | **stereo-EEG findings** | **Concordance of dMRI maps with [PET \| stereo-EEG]** | | **Operation** | **Histology** | **Outcome** |
| --- | --- | --- | --- | --- | --- | --- | --- | --- | --- | --- |
|  | **Patients with extratemporal TLE (Supplementary Fig. 24)** | | | | | | | | | |
| 01 | 10–14; M | 120 | subtle FLAIR abnormality in the mesial precentral sulcus extending anteriorly along the superior frontal sulcus | left frontal MFG abnormality | SOZ in mesial precentral sulcus | | No \| Yes | left frontal lesionectomy | focal gliosis | NSF |
| 02 | 15–19; M | 122 | ill-defined area of cortical and subcortical scarring involving the left parietal lobe in the region of the paracentral lobule. | reduced tracer uptake in the left superior parietal lobe (also in right supramarginal gyrus and left central sulcus) | not performed | | Yes \| n/a | left parietal lesionectomy | gliosis | SF |
| 03 | 10–14; F | 102 | MRI negative | asymmetric hypometabolism within the right temporal lobe | SOZ in parietal and temporal operculum with involvement of anterior long gyrus of insula | | Yes \| Yes | right parietal lesionectomy | non-specific changes | NSF |
| 04 | 5–9; M | 77 | Antenatal left MCA infarct and b/l white matter scarring (L>R); dilation of left lateral ventricle; atrophy of left caudate nucleus | Findings not marked; possible left hemispheric onset, most likely within left frontal temporal region | Left parieto-occipital onset | | No \| Yes | TPO offered but parents declined | n/a | NSF |
| 05^^^ | 10–14; M | 122 | Blurring of left Heschl's gyrus superior to insula | decreased uptake left insula with extension to pre/post central gyrus | SOZ posterior insula | | Yes \| Yes | thermocoagulation at time of stereo-EEG, subsequently x2 LITT of residual left insula lesion | n/a | SF |
| 06 | 5–9; F | 80 | subtle left hemispheric volume loss | left hemispheric onset, hypometabolism most marked in left central region contiguous in left superior parietal and left anterior/lateral temporal cortex | broad involvement of SMA and motor areas; SOZ not found | | Yes \| Yes | not offered | n/a | n/a |
| 07 | 10–14; F | 123 | left hemisphere smaller than right; white matter signal abnormalities most marked in left parietal lobe | left anterior parietal lobe hypometabolism extending throughout postcentral and supramarginal gyri, also lateral sulcus and superior left angular gyrus | seizure onset in the left posterior region with variable onset in superior, parietal or inferior temporal regions | | Yes \| Yes | Left TPO | FCD IIb | SF |
| 08 | 10–14; F | 123 | MRI negative | hypometabolism left subcentral gyrus and posterior insula | SOZ not identified; suspected involvement of left frontal-orbital frontal cortex but imaging not concordant | | Yes \| No | VNS offered; out of area | n/a | n/a |
| 09 | 10–14; F | 30 | no definite lesion but: Right superior frontal gyrus shows variant transversely oriented sulcation anteriorly with mild gyral expansion and blurring of grey-white junction. | suggestive of right frontal onset, probably within the MFG | widespread irritative zone with electrographic seizures in MFG, SFG and orbitofrontal region; SOZ not found | | Yes \| Yes | VNS | n/a | NSF |
| 10 | 10–14; F | 31 | MRI negative | left hemispheric focus most likely in left MFG, alternatively left anterior temporal lobe | widespread involvement of the left frontal lobe is seen at seizure onset; a focal seizure onset could not be identified. | | Yes \| Yes | Not surgical candidate | n/a | n/a |
|  | **Patients with TLE (Supplementary Fig. 25)** | | | | | | | | | |
| 11 | 15–19; M | 106 | MRI negative | Bilateral hypometabolism, left more than right, involving left parietal and posterior insula | Wide left hemispheric onset; left hippocampal after-discharges | | Yes \| Yes | Left temporal lobectomy and amygdalohippocampectomy | HS (subtle) | NSF |
| 12 | 10–14; M | 110 | Ill-defined loss of grey-white matter differentiation in the left temporal lobe | Left temporal hypometabolism | Not performed | | Yes \| n/a | Left temporal lobectomy | DNET (WHO 1) | SF |
| 13 | 5–9; F | 61 | MRI negative | Bilateral temporal hypometabolism, more pronounced right posterior temporal | Not performed | | Yes \| n/a | Right temporal lobectomy | HS | NSF |
| 14 | 10–14; F | 120 | Impression of subtle blurring right insular cortex and adjacent frontal operculum anteriorly | Multifocal hypometabolism, more pronounced right cerebral hemisphere | Right posterior insula | | Yes \| Yes | Right LITT (at time of stereo-EEG) | n/a | NSF |
| 15 | 15–19; M | 204 | MRI negative | Left temporo-insular opercular hypometabolism, subtle peri-rolandic | Posterior temporal and parietal region; unique focal onset not found | | Partial (peri-rolandic and contralateral) \| Partial (left temporal only) | Not performed | n/a | N/A |

Supplementary Table 1 **Demographic and clinical characteristics of the patients with MRI-negative temporal and extra-temporal epilepsy with PET images for comparison**. ^ same patient as patient 18. DNET; Dysembryoplastic Neuroepithelial Tumour, HS; hippocampal sclerosis, LITT; laser interstitial thermal therapy, MCA; middle cerebral artery, MFG; middle frontal gyrus, SFG; superior frontal gyrus, SMA; supplementary motor area, TPO, temporo-parieto-occipital, VNS; vagus nerve stimulation.

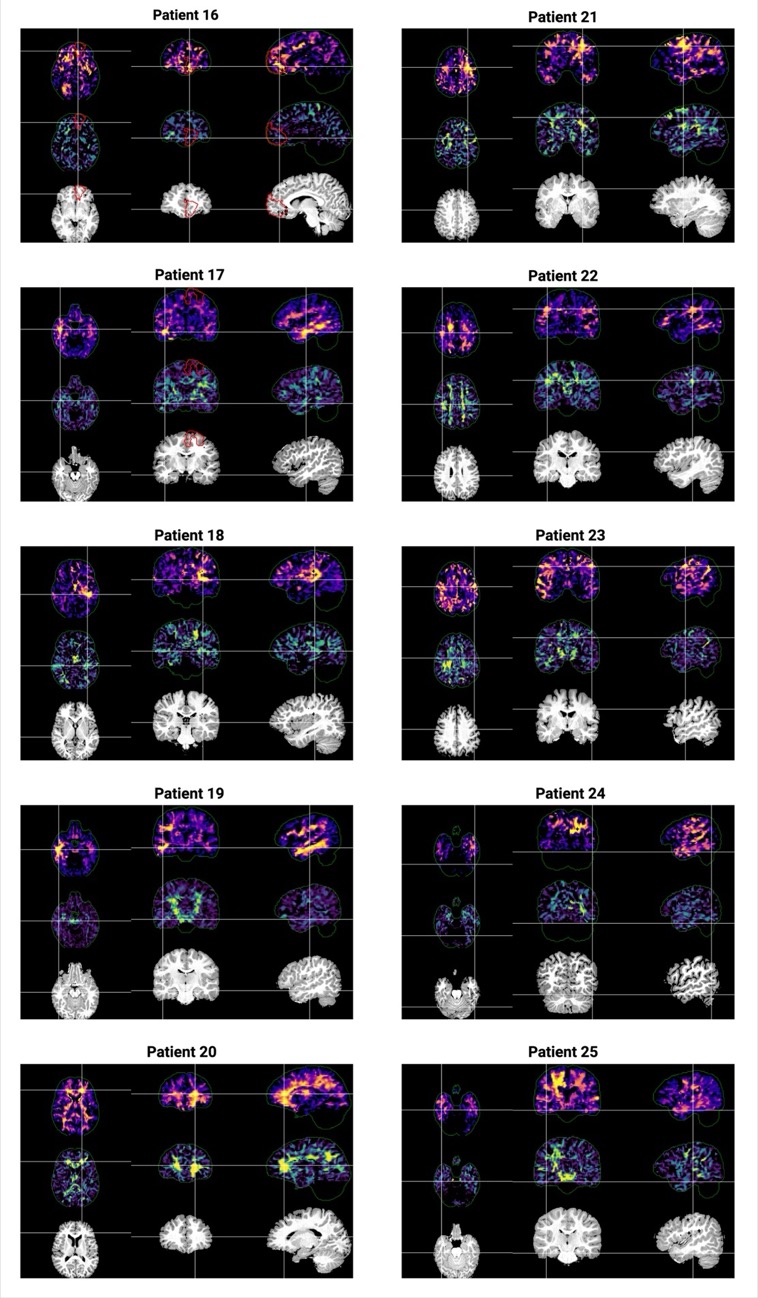

Supplementary figure 11 Per-patient normative diffusion abnormalities in radiological orientation for MRI lesion-negative patients with a confirmed or suspected extratemporal SOZ, who did not have PET images but had PET reports available. Top panel: unthresholded TFCE normative MD map (using positive z-values, reflecting elevated diffusivity relative to controls). Middle panel: unthresholded TFCE normative FA map (using negative z-values, reflecting reduced anisotropy relative to controls). Bottom panel: Linearly aligned T1 scan in diffusion template space. Black crosshairs: SOZ-designated stereo-EEG contacts (where applicable). Red outline: resection masks (where applicable)

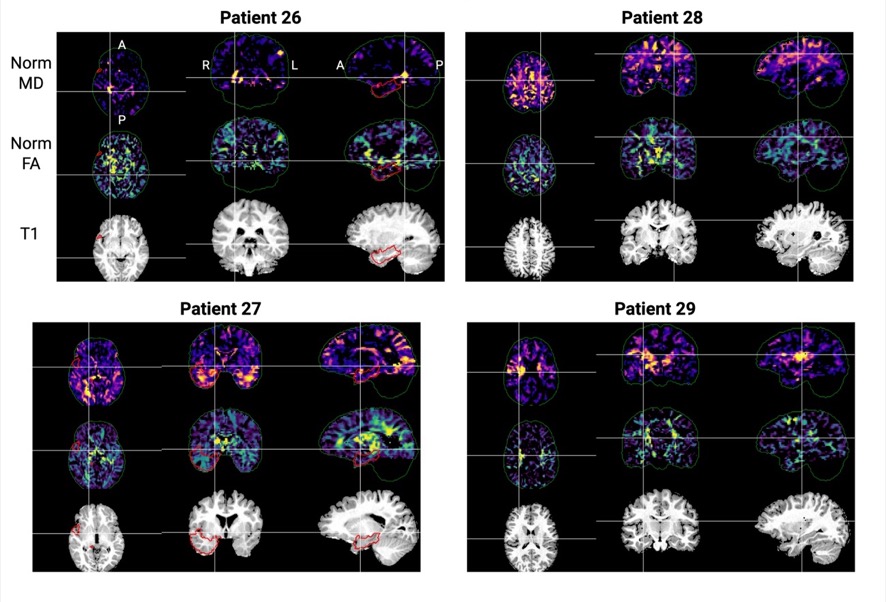

Supplementary figure 12 Per-patient normative diffusion abnormalities in radiological orientation for MRI lesion-negative patients with a confirmed or suspected temporal SOZ, who did not have PET images but had PET reports available. Top panel: unthresholded TFCE normative MD map (using positive z-values, reflecting elevated diffusivity relative to controls). Middle panel: unthresholded TFCE normative FA map (using negative z-values, reflecting reduced anisotropy relative to controls). Bottom panel: Linearly aligned T1 scan in diffusion template space. Black crosshairs: SOZ-designated stereo-EEG contacts (where applicable). Red outline: resection masks (where applicable)

| **Patient** | | **Age range; sex** | **Duration of epilepsy (months)** | **MRI diagnosis** | **PET** | **stereo-EEG findings** | **Spatial concordance of dMRI maps with [PET \| stereo-EEG]** | **Operation** | **Histology** | **Outcome** |
| --- | --- | --- | --- | --- | --- | --- | --- | --- | --- | --- |
|  | **Patients with extratemporal TLE (Supplementary Fig. 26)** | | | | | | | | | |
| 16 | | 15–19; M | 40 | MRI negative | Reduced uptake right medial superior frontal gyrus and left superior medial frontal gyrus | SOZ in left orbitofrontal region | Partial (also c/l orbitofrontal abn.) \| Partial (also c/l orbitofrontal abn.) | left frontal lesionectomy | mild gliosis | NSF |
| 17 | | 5–9; M | 64 | MRI negative (processed MP2RAGE possible subtle blurring anterior cingulate and left precuneus) | Hypometabolism in left mesial frontal region and right operculum/temporal lobe | SOZ from left posterior SFG with quick involvement of motor/face regions and posterior SFG | Right-sided findings only \| No | thermocoagulation at time of stereo-EEG then left frontal lesionectomy | FCD IIb | SF |
| 18^^^ | | 10–14; M | 117 | MRI negative | decreased uptake left insula with extension to pre/post central gyrus | SOZ posterior insula but spatial extent uncertain | Yes \| Yes | thermocoagulation at time of stereo-EEG | n/a | NSF |
| 19 | | 10–14; M | 138 | Subtle blurring of grey/white matter junction in right frontal and temporal regions | inconclusive: bilateral focal areas of reduced FDG uptake, possibly representing multifocal epilepsy of ischaemic changes | wide network in right frontal, posterior temporal and parietal regions; SOZ not found | Yes \| Yes | VNS | n/a | NSF |
| 20^*^ | | 10–14; M | 34 | MRI negative | Hypometabolism in left inferior frontal region/posterior insula/posterior frontal operculum | left frontal eye field | Yes \| Yes | Thermocoagulation at time of stereo-EEG | n/a | NSF |
| 21 | | 15–19; F | 106 | MRI negative | diffuse left hemispheric hypometabolism | SOZ not found; multiple seizure types with wide network in left frontal region and additionally left temporal region | Yes \| Yes | VNS offered; out of area | n/a | n/a |
| 22^*^ | | 10–14; M | 36 | MRI negative | mild hypometabolism in right central and temporal regions | SOZ not identified; possible parietal origin | Partial (central) \| No | n/a | n/a | n/a |
| 23 | | 15–19; M | 62 | MRI negative | possible hypometabolism left post central area | SOZ not identified | Partial (also c/l changes) \| n/a | VNS | n/a | NSF |
| 24 | | 15–19; F | 69 | MRI negative | left temporal, potentially mesial origin | SoZ around precuneus /posterior cingulate around thermocoagulation of previous stereo-EEG following which seizure free for 8 months - abnormal sulcus in this area | Yes \| Yes | RFTC at time of second stereo-EEG | n/a | n/a |
| 25 | | 5–9; F | 17 | subtle blurring of grey-white matter in right anterior frontal lobe | mild asymmetrical hypometabolism in the mesial right temporal lobe extending into the temporal pole | Repeat SOZ not found; diffuse left hemispheric network | Yes \| No | Thermocoagulation at time of stereo-EEG, then VNS; subsequently removed as ineffective | n/a | NSF |
|  | **Patients with temporal TLE (Supplementary Fig. 27)** | | | | | | | | | |
| 26 | | 10–14; M | 42 | MRI negative | Right antero-mesial temporal pole hypometabolism | Not performed | Partial (posterior hippocampus) \| n/a | Right temporal lobectomy and amygdalohippocampectomy | HS | NSF |
| 27 | | 5–9; M | 20 | MRI negative | Bilateral temporal hypometabolism | Right hippocampus | Yes \| Yes | Right temporal lobectomy | Non-diagnostic | SF |
| 28 | | 15–19; M | 207 | Diffuse subtle loss of left hemispheric volume concordant | Normal | Widespread onset including mesial frontal, frontal operculum, orbitofrontal, precuneus; no single seizure onset identified | n/a \| partial (FA>MD) | Planned VNS but lost to follow up after relocation | n/a | Not available |
| 29 | | 15–19; F | 195 | Subtle asymmetry of right insular cortex | Right posterior frontal and parietal hypometabolism; bilateral temporal hypometabolism right more than left | Focal onset from right posterior insula | Yes \| Yes | Right LITT (at time of stereo-EEG) | n/a | NSF |

Supplementary Table 2 Demographic and clinical characteristics of the patients with MRI-negative epilepsy without PET images for comparison (PET reports only). ^ same patient as patient 05. * Pertain to the same patient. HS; hippocampal sclerosis, LITT; laser interstitial thermal therapy, RFTC; radiofrequency thermal coagulation, SFG; superior frontal gyrus, SOZ; seizure onset zone, VNS; vagus nerve stimulation.

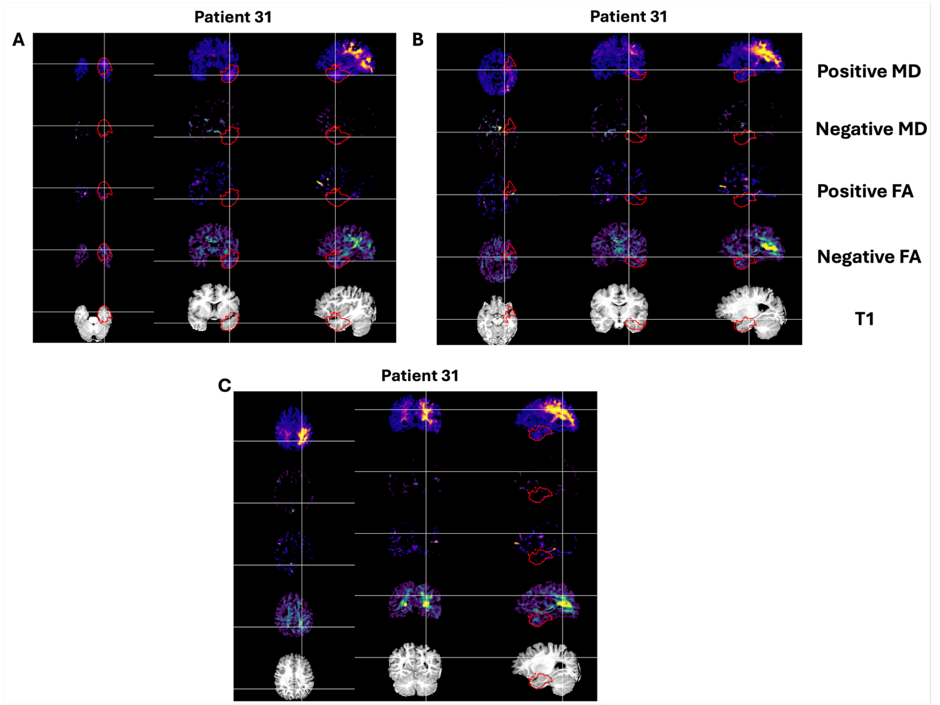

Supplementary figure 13 Positive and negative normative MD and FA modalities for patient 31 **(A)** Centred on the left temporal resection **(B)** Centred in the left mesial temporal region. (**C)** Centred on the left parieto-occipital abnormality. This case was a child with focal epilepsy beginning in the neonatal period, initially associated with hypoglycaemia. Preoperative seizures involved headache or a “busy bee” sensation, anxiety or fear, with partial awareness and preserved speech. Scalp EEG, seizure semiology and SPECT were concordant for a left temporal epileptogenic focus, although MRI also showed broader bilateral posterior abnormalities, more marked on the left, including periventricular signal change/volume loss near the occipital horns, underdevelopment of white matter, especially in the parietal and occipital lobes, more pronounced on the left, and a smaller, high-signal left hippocampus consistent with mesial temporal sclerosis. Language fMRI was difficult to interpret, with some right temporal activation, and the patient was partially sighted with developmental delay of approximately 18 months. Following multidisciplinary discussion, a left anterior temporal lobe resection and amygdalohippocampectomy was performed, rather than a more posterior resection, because of concern about language and visual-field risks and uncertainty regarding the contribution of posterior abnormalities. Histopathology confirmed hippocampal sclerosis. Postoperatively, seizures increased. The patient was later offered stereo-EEG with a view to possible left temporal-parietal-occipital surgery/disconnection, but this was declined by the parents. Over time, the patient developed drop seizures, and later video telemetry demonstrated a new contralateral right nocturnal tonic seizure onset, after which he was no longer considered a candidate for stereo-EEG or left TPO surgery.

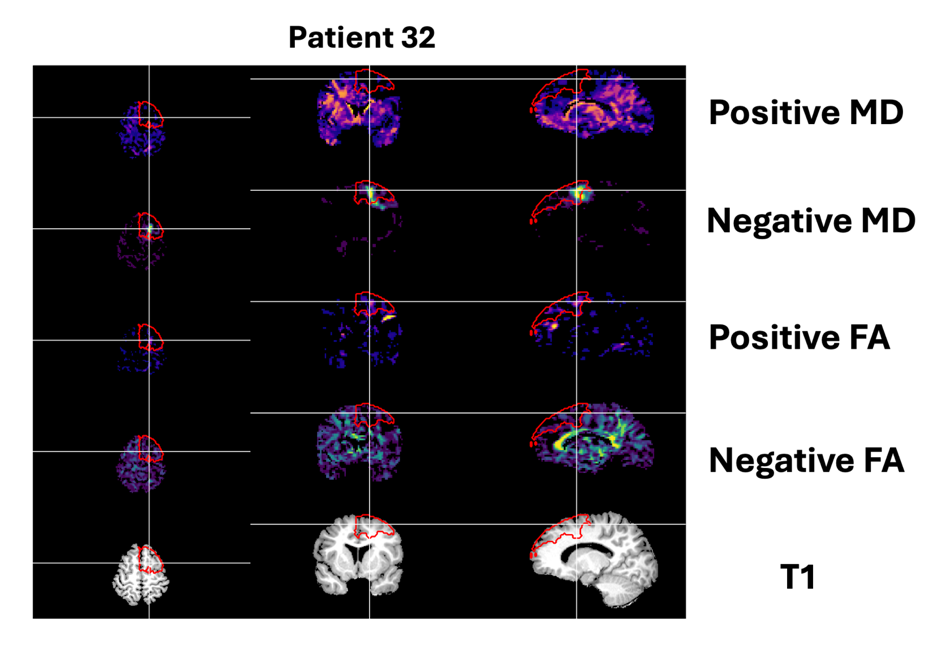

Supplementary figure 14 Positive and negative normative MD and FA modalities for patient 32 **(A)** Centred on the left frontal lesionectomy (red outline).

| **Cohort** | **Reason for exclusion** | **Number** |
| --- | --- | --- |
| Resection | Age | 62 |
|  | Different scanner and/or scanning protocol | 50 |
|  | Missing scans | 38 |
|  | Previous operation | 9 |
|  | Tuberous sclerosis complex | 7 |
|  | Hemispherotomy | 2 |
|  | Insufficient follow-up | 1 |
|  | Missing histology result | 1 |
|  | Poor preprocessing | 1 |
|  | Duplicate | 1 |
| Stereo-EEG | Age | 23 |
|  | Previous resection | 14 |
|  | Missing clinical information | 13 |
|  | Tuberous sclerosis complex | 13 |
|  | Different scanner and/or scanning protocol | 7 |
|  | Missing scans | 6 |
|  | Poor preprocessing | 3 |
|  | Age and TSC | 2 |
|  | Age and previous resection | 1 |
|  | Subdural grid | 1 |

Supplementary Table 3 Number of patients excluded across the cohorts and reasons for exclusion

**A:**
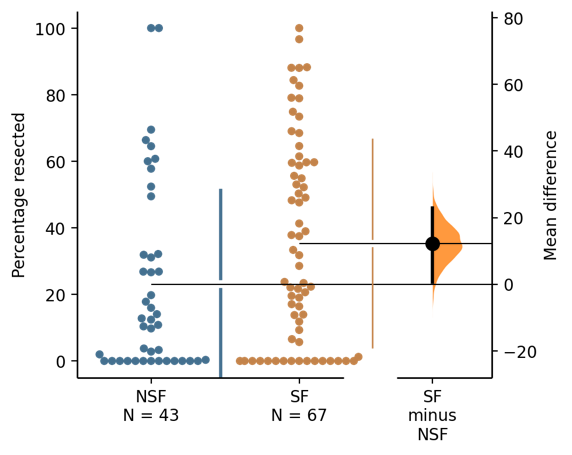
**B:**
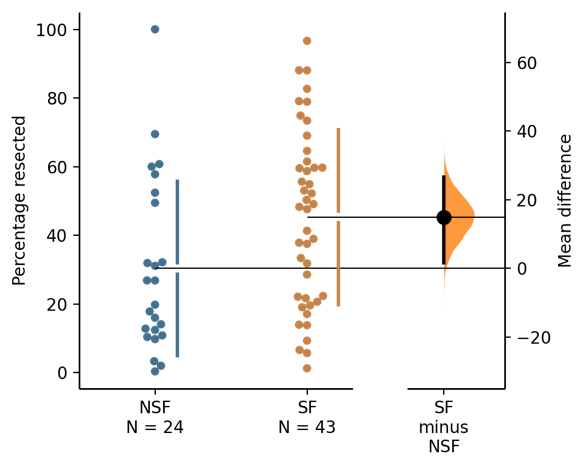

**Supplementary Figure 15 Extent of resection of abnormal diffusion MD clusters (derived using a z>3 threshold only) and association with seizure freedom. (A)** Percentage resected of the dominant MD-derived cluster for all patients, including those with none of the dominant cluster resected. The unpaired mean difference between NSF and SF is 12.2 [95.0%CI 0.559, 22.9]. The P value of the two-sided permutation t-test is 0.041. **(B)** Percentage resected of the dominant MD-derived cluster for patients with >0% resection. The unpaired mean difference between NSF and SF is 15.0 [95.0%CI 1.66, 26.7]. The P value of the two-sided permutation t-test is 0.0244.
